# Media-driven adaptive behavior in pandemic modeling and data analysis

**DOI:** 10.1101/2024.04.17.24305855

**Authors:** Zhiyuan Yu, David Gurarie, Qimin Huang

## Abstract

Human behavior and public attitudes towards preventive control measures, such as personal protection, screening, isolation, and vaccine acceptance, play a crucial role in shaping the course of a pandemic. These attitudes and behaviors are often influenced by various information sources, most prominently by social media platforms.

The primary information usually comes from government bodies, e.g. CDC, responsible for public health mandates. However, social media can amplify, modify, or distort this information in numerous ways. The dual nature of social media can either raise awareness and encourage protective behaviors and reduce transmission, or have the opposite effect by spreading misinformation and fostering non-compliance.

To analyze the interplay between these components, we have developed a coupled SIR-type dynamical model that integrates three essential components: (i) disease spread, as reported by official sources; (ii) the response of social media to this information; and (iii) the subsequent modification of human behavior, which directly impacts the spread of disease.

To calibrate and validate our model, we utilized available data sources on the Covid-19 pandemic from a one-year period (2021-2022) in the United States, as well as data on social media responses, particularly tweets. By analyzing the data and conducting model simulations, we have identified significant inputs and parameters, such as initial compliance levels and behavioral transition rates. These factors enable a quantitative assessment of their contributions to disease outcomes, including cumulative outbreak size and its dynamic trajectory.

This modeling approach gives some valuable insights into the relationship between public attitudes, information dissemination, and their impact on the progression of the pandemic. By understanding these dynamics, we can inform policy decisions, public health campaigns, and interventions to effectively combat the spread of Covid-like pathogens and future pandemics.

## 2 Introduction

Infectious diseases posed a significant threat throughout human history. Predicting their emergence, progression, and response stands as a paramount societal goal. Infections are categorized by transmissibility and geographic scope [1], and the 21st century has witnessed several pandemics: severe acute respiratory syndrome (SARS) (2002-2003), Swine flu (2009-2010), Middle East respiratory syndrome (MERS) (2015-2023), and COVID-19 (2019-present) [2].

Multiple factors can contribute to disease spread. For vector-borne diseases they include ecology and environment; for communicable diseases population makeup and behavior (social interactions) play paramount roles.

Behavioral adaptation combines risk assessment from perceptual cues, and the appropriate preemptive or mitigating responce. [3, 4]. Empirical evidence shows the potential of behavioral adaptation to curtail infection, via reduced social mixing, and other protective practices like face-masking, hand-washing, public space decongestion, surface sanitization, social distancing, and quarantine. They played crucial role in containing outbreaks like SARS, H1N1 (Swine flu), and Covid-19 [5–9].

Behavioral shifts are categorized as reactive or proactive [4]. Amid the pandemic, perceptual cues can come from different sources, like social media and peer interactions. Some professional groups (e.g., healthcare workers) undertake preventive measures regardless of immediate risk. Today, social media serves as source of information, and a potent driver of behavior. On the one hand, it could amplify awareness and encourage preventative actions, as well as serving the tool for disseminating health information and government containment policies [10, 11], On the other hand, it creates a platform for propagation of rumors and misinformation, which impede health-conscious decisions, and foster mistrust in governmental bodies, policies and scientific expertise [12].

To explore the interplay between behavior, information, and disease, a number of modeling approaches were developed based on game theory, network modeling, individual-based (agewnt) approach, and compartmental SIR models (suscepible-infected-recovered) [13]. Behavior can enter such models in different forms, e.g. via prescribed behavioral strata and switching patterns, by adaptively changing model parameters, or social network topology. A number of recent and earlier works employed network methodology to explore behavior-disease dynamics, and address some basic theoretical questions [14–23]. Although network-based models can provide deeper intuition on theoretical level, they are hard to train and fit to a particular dataset. Other work Funk et al. (2010), Misra et al. (2011), Samanta et al. (2013), Misra et al. (2015), and Agaba et al. (2017) employed SIR formulation for two dynamic behavioral strata (aware, unaware), and in their assumptions, aware hosts have lower contact rate than the unaware hosts [24–28]. However, Funk et al. and Agaba et al. used direct contact as the main driver of behavioral switching, while Misra et al. and Samanta et al. used the pandemic-stimulated campaigns as the driver of behavioral switching. To the best of our knowledge, Misra et al. (2011) were the first who included the media-driven behavioral change in their SI model, and they chose the total infected population size as the driver of social media. The model proposed by Misra et al. is interpretable and exhibits interesting dynamics, but it lacks some critical aspects, such as the recovered compartment and the aware infective population. In addition, they used a first-order decay term, independent of social media, to model the loss of awareness, which is unrealistic as aware hosts can also be affected by social media and maintain their awareness [25]. Rai et al. (2022) and Koutou et al. (2023) incorporated social media into their models, but in their setups, media inputs mainly affect the instantaneous infection rate, and there are no clear behavioral stratifications in their models [29, 30]. Guo et al. (2021) explicitly included media reports of cases as a modulating variable for infection rate and quarantine rate, and they chose the hospitalized population size as the driver of social media [31]. Tiwari et al. (2021) stratified the susceptible pool into three behavioral compartments (unaware, highly active aware, and loss active aware), and they let social media drive hosts into two aware pools. However, they didn’t apply the behavioral stratification to other compartments, such as the infective population and the recovered population. Also, Tiwari et al. still used the first-order awareness decay, which as mentioned before, is not realistic [32].

In this study, our goal is to develop a simple and robust behavior-modified ODE model to validate the importance of behavior and media in disease transmission, and we want to explore the effects of behavior and media on the pandemic outcomes. Our setup is close to the combination of the setups of Funk et al. (2010) and Misra et al. (2011), but we explicitly used pandemic-stimulated social media as the driver of behavioral switching. Also, unlike previous works, our aware/unaware (compliment/noncompliant) behavioral pools are driven by ‘media attention’, which in turn feeds on the state of the pandemic. Such adaptive behaviors, in particular acceptance of vaccines, modulate transmission rate and determine the course of the pandemic.

## 3 Method

### 3.1 Modeling setup

We constructed the model based on the classical SIR model, and we stratified each epidemiological compartment into two behavioral strata. As shown in Table 1, our model consists of seven variables in total. Same as the classical SIR model, *S*, *I*, and *R* denote susceptible, infected, and recovered hosts. In addition, each epidemiological compartment has two behavioral strata: non-compliant (subscript *N*) and compliant (subscript *C*). Finally, *M* represents the effective pandemic-related media, and here “effective” means that the media is accessible and frequently viewed on the social media platform.

**Table 1:** Variable table.

| Variable | Description |
| --- | --- |
| $S_N$ | Noncompliant susceptible hosts |
| $S_C$ | Compliant susceptible hosts |
| $I_N$ | Noncompliant infected hosts |
| $I_C$ | Compliant infected hosts |
| $R_N$ | Noncompliant recovered hosts |
| $R_C$ | Compliant recovered hosts |
| $M$ | Effective Pandemic-related media |

To recap, the studies conducted prior to this one have demonstrated certain limitations in their actions and media inputs, such as the absence of a consistent behavioral classification for all epidemiological compartments, unfinished behavioral transition, and media drivers that are sensible but yet to be validated. However, media input plays a key role in human behavior, which is a key part in epidemiological models for pandemic. Considering these challenges, we are proposing a straightforward and easy-to-understand framework for examining the impact of behavior and media inputs on the spread of infectious diseases. By combining data analysis methods with theoretical modeling, we are not only able to validate our model framework using actual data but also provide reliable inferences from that data. We believe that our work can offer more insights into the integration of behavior and media into epidemiological models.

We summarized our ideas in Figure 1. First, we included two dynamic behavioral strata for all three epidemiological compartments (susceptible, infective, and recovered), designated as compliant (C) and non-compliant (NC). We assumed that compliant hosts (C) have reduced disease exposure and transmission, and they accept vaccination. In contrast, non-compliant pool (NC) has higher exposure - transmission rates relative to (C), and reject vaccination. Besides behavioral pools, our model contains the dynamic social media compartment - its disease-related content, that get input from the current epidemic state, and serves as a principal driver of behavioral change (NC to C transitions). We assumed that social media serves as the major massive control of hosts’ behavior, and its effect surpasses any other behavioral impact factors. We will use data analytic strategies to justify this choice. Our media divers are different from those of the previous works as we think social media usually respond to the change of the pandemic state (e.g. new incidence in a week, which is the information we typically saw on social media during the Covid-19 pandemic) rather than the pandemic state itself. In addition, the change in the pandemic state is usually more observable and changes more quickly than the pandemic state itself, so the change in the pandemic state may be better in explaining the short-term fluctuation in social media.

**Figure 1:**
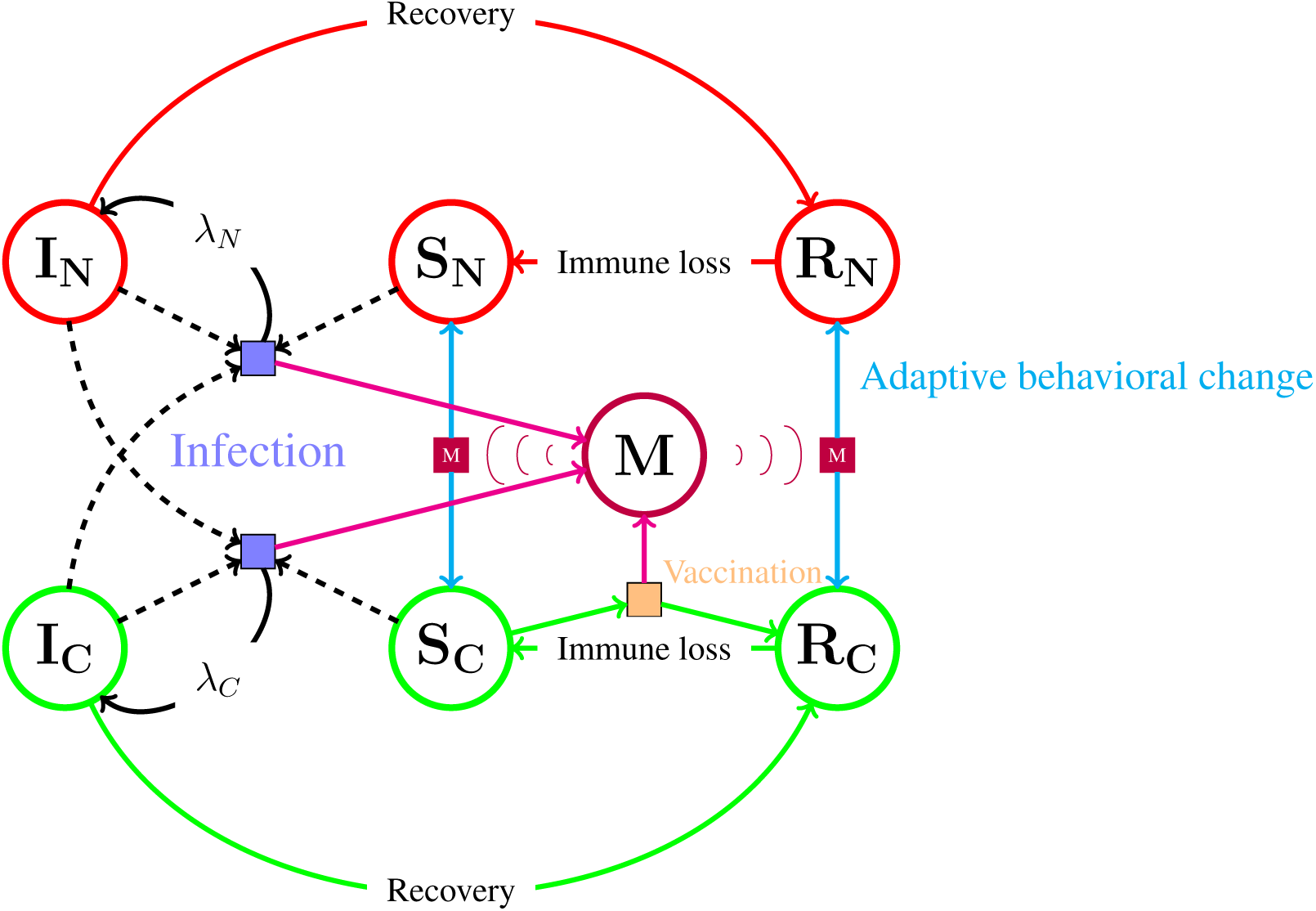
Schemetic diagram of the coupled-SIR model. Every epidemic compartment has two behavioral strata: compliant (green) and non-compliant (red). The left part of the diagram shows the cross-infection between two infective behavioral strata and two susceptible behavioral strata, where being compliant can reduce disease transmission and exposure. The right part of the diagram shows the behavioral switching driven by the quantity of pandemic-related media, and the state of the pandemic modulates the quantity of media. From our later analyses, both the content and the quantity of the media exhibit strong associations with infection and vaccination, so they serve as the primary driving factor of social media, as shown by the pink arrows in this diagram. The recovery and immune loss happen independently in each behavioral pool.

We assumed that two behavioral pools change dynamically throughout the pandemic, but we ignored the direct behavioral transition between compliant infective and noncompliant infective due to the short infectious period. Also, we assumed that hosts’ behavioral switching pattern stays constant throughout time, meaning that host behavior will be driven by the perceived state of infection and vaccination via social media. By assuming that disease-related media contains a roughly fixed proportion of appropriate content that can make hosts compliant, we used the total amount of social media rather than its content as the driver of the compliance switching, and we will justify this assumption with text mining and sentimental analysis later. Specifically, noncompliant hosts are more likely to become compliant after seeing more disease-related media, and compliant hosts will keep paying attention to media and thereby maintain their compliance. Thus, the net-behavior-flow goes from non-compliant to compliant when there is a lot of disease-related social media, and the flow reverses its direction when social media decreases. Moreover, we assumed that hosts are more likely to lose compliance after getting recovered, meaning that there is a negative feedback loop: when social media increases due to infection or vaccination, more hosts will take the vaccine and recover, leading to loss of average compliance and subsequently less vaccination, which slows down social media production. One more assumption for social media is the turnover because newly created media can replace the old media, and other media can replace disease-related media when the rate of producing the disease-related media decreases.

The choice of infection and vaccination rates as the prime drivers of social media, as shown by the magenta arrows in Figure 1, was not arbitrary, but derived from data analysis via machine learning tools.

Table 2 shows all nine parameters in our model. Parameter *r* is the basic disease transmission rate, or the approximate average number of infections per infected individual given that almost all the individuals in the population are susceptible, between an arbitrary infected individual and an arbitrary susceptible individual. Parameter *ω* is a number between 0 and 1, and it represents the reduced transmission/exposure for being compliant relative to being non-compliant. In addition, the per-pair disease transmission rate depends on the identities of the disease carrier (I) and the disease recipient (S). Therefore, the pair *I_N_* &*S_N_*, *I_C_*&*S_N_*, *I_N_* &*S_C_*, and *I_C_*&*S_C_* have the transmission rate 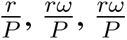, and 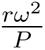, where *P* is the total population size. Parameter *A* is the maximum per capita behavior switching rate, and the per capita behavior switching rate converges to *A* when the amount of media *M* approaches infinity. Parameter *M_T_* is the compliance threshold, and when *M_T_* gets larger, hosts need more disease-related media to become compliant. Parameters *γ* and *δ* are the recovery rate and immune loss rate. Parameter *v_C_* is the vaccination rate of the compliant individual, and we assume only *S_C_* takes the vaccine for simplicity. Parameter *ɛ* is the media due to new cases, which corresponds to the media such as tweets that report the incidence or other incidence-related information. Parameter *σ* is the media due to new vaccination cases, which corresponds to the media that reports the news about new vaccines, pharmaceutical companies, and cases of vaccinations.

**Table 2:** Parameter table.

| Parameter | Description | Unit |
| --- | --- | --- |
| $r$ | Basic transmission rate | week<sup>-1</sup> |
| $\omega$ | Reduced transmission/exposure for being compliant relative to being non-compliant | Unitless |
| $A$ | Maximum behavioral switching rate | week<sup>-1</sup> |
| $M_T$ | Compliance threshold | Unitless |
| $\gamma$ | Disease recovery rate | week<sup>-1</sup> |
| $\delta$ | Immune loss rate | week<sup>-1</sup> |
| $v_C$ | Per capita vaccination rate by the compliant pool | week<sup>-1</sup> |
| $\epsilon$ | Media due to new cases (incidence) | Unitless |
| $\sigma$ | Media due to new vaccination cases | Unitless |

One full model (equation 1) consists of the classical infection-recovery-wanning-reinfection cycle and the adaptive behavioral switching. Term *ϕ* denotes the switching between non-compliant and compliant strata, and we modified the equation such that the total behavioral switching rate between one non-compliant individual (S or R) and one compliant individual (S or R) *ϕ_N→C_* + *ϕ_C→N_* is *A*, corresponding to our assumption that hosts have constant behavioral rigidity throughout the time. In addition, we made the rate of change of disease-related media 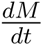 linearly dependent on the total infection rate and the vaccination rate with an additional media turnover term, and we justified this form with our data analysis results. Our model focuses on population dynamics and large-scale behavioral control via media, so it becomes more accurate when the population size is sufficiently large.

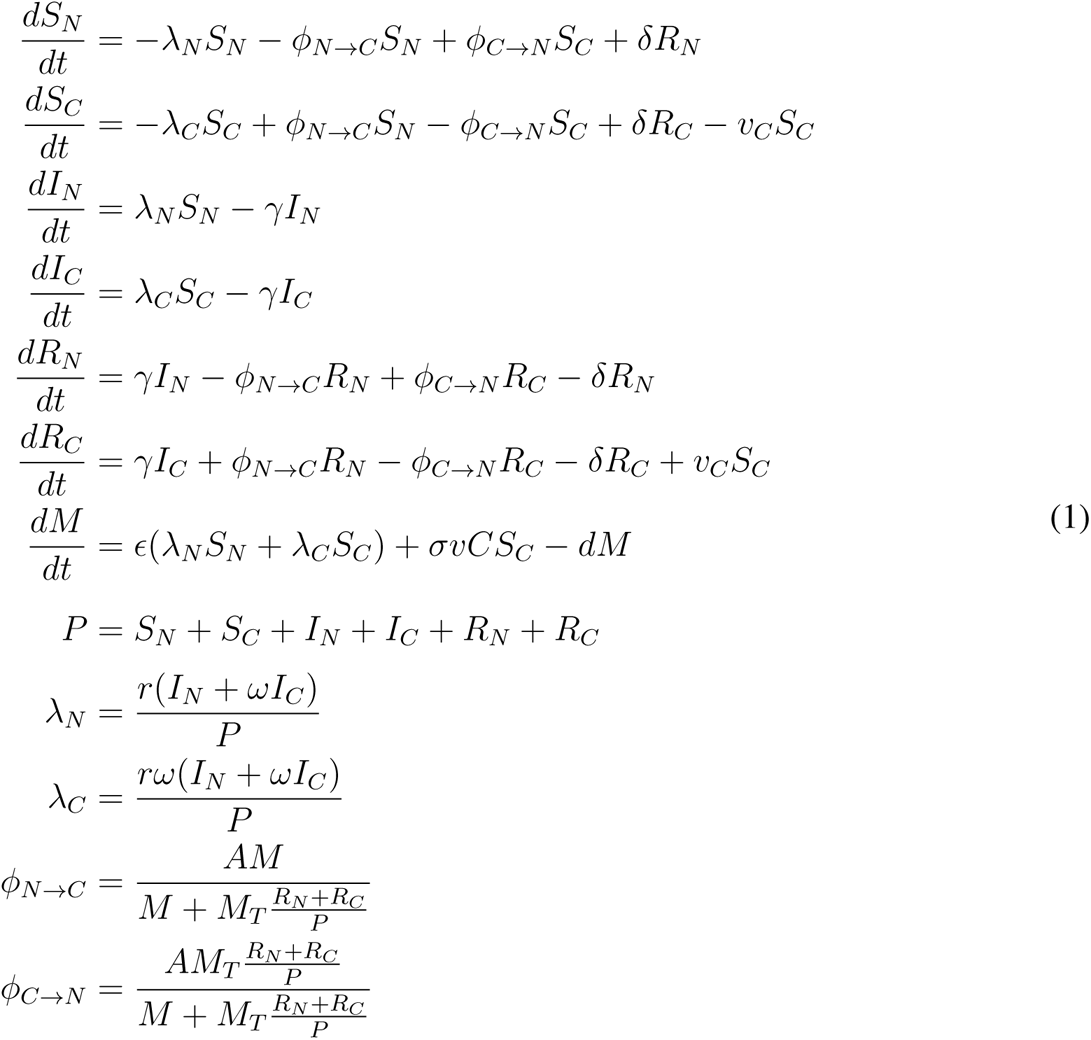

### 3.2 Data sources and analysis

#### 3.2.1 Data Description

In order to model and validate behavior and media responses effectively, we opted to utilize US Covid-19 data. This choice is motivated by several key factors. Firstly, Covid-19 stands as the most recent widespread global epidemic up until 2023. The collective endeavors of society, institutions, and governments have resulted in comprehensive Covid-19 data availability. This data holds significant value for both research and general applications, including media content creation. Secondly, the Covid-19 pandemic has witnessed a distinct increase in individuals’ accessibility to social media, unlike any prior pandemics. This surge is attributed to advancements in hardware, software, and heightened utilization of social media platforms. Consequently, individuals have engaged with social media more extensively throughout the Covid-19 outbreak. Thirdly, our data is sourced from https://ourworldindata.org, a reputable and widely acknowledged platform catering to diverse audiences—ranging from researchers and journalists to policymakers. Remarkably, this platform’s data is cited in excess of 50,000 media articles, including over 20,000 contributions from prominent outlets like the New York Times and BBC. This broad journalistic accessibility underscores the data’s pivotal role in capturing the direct influence of the pandemic on social media.

For our primary Covid-19-related media source, we singled out Twitter, considering its prominence and representative nature of Covid-19-related social media activity. We procured Covid-19-associated tweet IDs from Chen et al., who have curated such data since January 2020 [33]. By merging the Covid-19 pandemic dataset with Twitter data, we obtained a comprehensive dataset encompassing both domains.

Figure 2 illustrates the selection and plotting of two time-series variables from Covid-19 pandemic and Twitter data, using epidemic weeks as time units. The data is divided into three segments: 2020, 2021, and 2022. The 2020 data is omitted due to high Covid-19 data missingness. Gaps in Twitter data collection, attributed to Twitter API restrictions by Chen’s group, are observed. For feature selection, model selection, and model calibration, the 2021 data is employed. Validation of our calibrated model is conducted using the 2022 data. Notably, the evolving coronavirus exhibits multiple strains taking turns as dominant. Figure 2 indicates prevalent variant timeframes. Since alpha and beta variants are somewhat similar to each other, we assumed that our model can mostly recover the patterns in the year 2021’s data with a single solution trajectory. However, since the omicron variant has shown several genetic and phenotypic variations that can make it escape the immunity established with respect to the alpha or the beta variant, we decided to reset the entire recovered pool to susceptible at the point when the omicron variant becomes prevalent.

**Figure 2:**
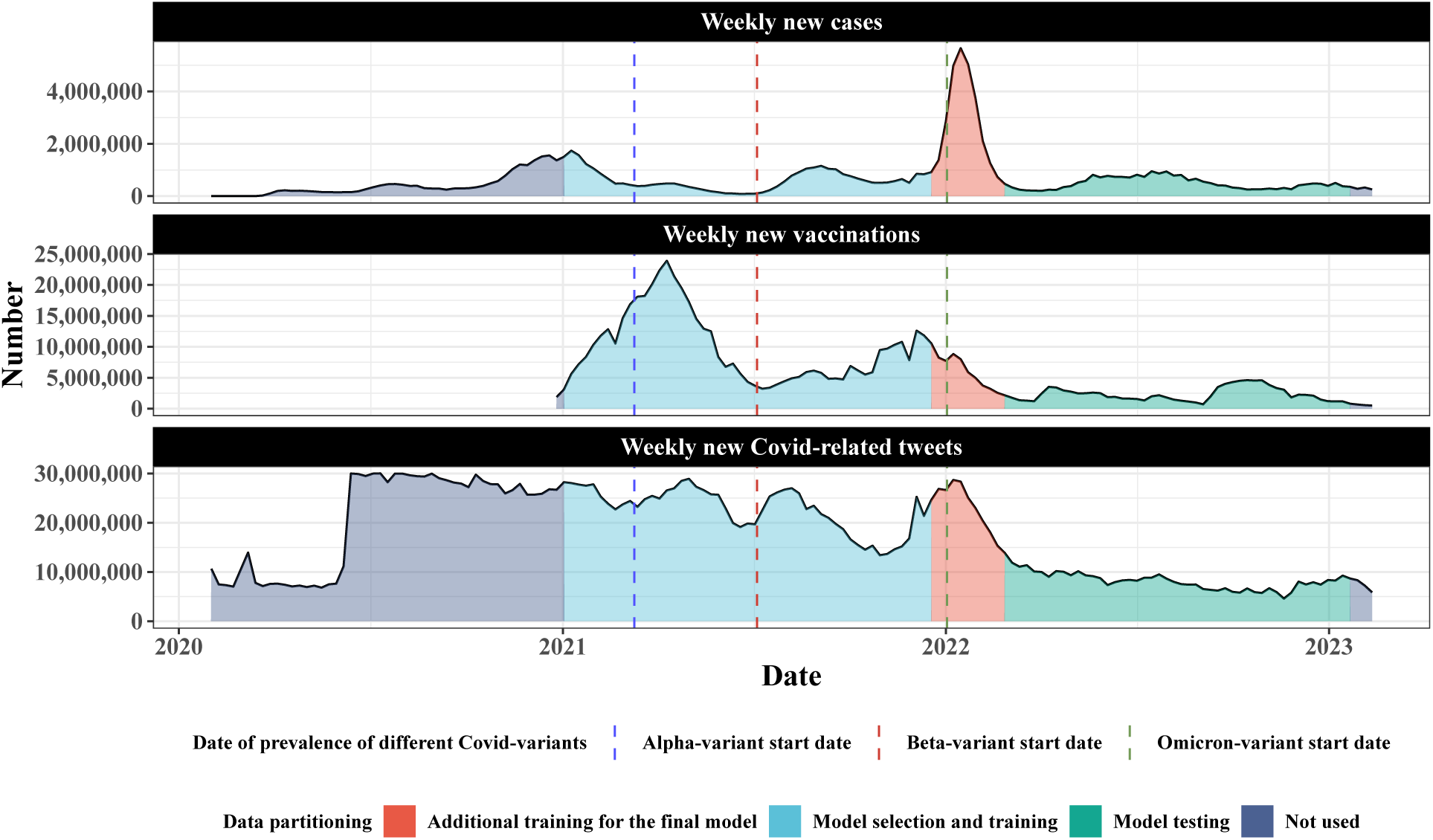
Combined time-series dataset including two variables from the pandemic data and Twitter data from the start of 2020 to the start of 2023. The dataset has three parts corresponding to their usage in this study: not used, model selection and training, and model testing. We discarded the “not used” part due to its high data missingness. Model selection includes the selection of the most predictive features and the functional form for the social media, and we used nested cross-validation for these tasks. Model training on 2021’s data is the Bayesian inference with respect to the final differential equation model. We finally tested the calibrated differential equation model on 2022’s data. We also marked the dates of the prevalence of different Covid variants.

As shown in Figure 1, we didn’t stratify the social media compartment, and one of our assumptions is that disease-related media contains a roughly fixed proportion of appropriate content that can make hosts compliant. To validate this assumption, we did text mining and sentimental analysis to quantify the proportion of appropriate content in disease-related media, as explained in the next section (3.2.2). We used the cleaned unigram and bigram data published by Banda et al. (2021), which can be acquired at https://github.com/thepanacealab/covid19 twitter [34]. The raw data contains Twitter ids related to Covid-19 posted every second since March 2020, and the cleaned data contains the top 1000 unigrams and bigrams each day. We used the cleaned data to get a general view of the content of disease-related tweets each day. Indeed, our subsequent text analytical results only reflect the general media content trend and composition, while in reality, social media’s content can vary between different geographical regions, different socioeconomic groups, and individual users.

#### 3.2.2 Text mining and sentimental analysis

Sentimental analysis is the process of analyzing the emotion behind each word, sentence, or larger text body. Specifically, we used sentimental analysis to see if disease-related media conveys the appropriate sentiments from hosts to hosts and raise hosts’ awareness of the pandemic in a positive manner. Indeed, here we assumed that the media with appropriate sentiments is generally the media with appropriate content to promote hosts’ compliance, but in reality, individuals have different sensitivities and responses to the same media content. There are many ways to conduct sentimental analyses, but most of them either utilize sentimental lexicons or machine learning models. Machine learning models such as SVM, deep learning, and other common classifiers are able to classify large text bodies based on the context and are scalable to large data volumes, but these models typically require careful calibration and tuning [35] to achieve good accuracy. Lexicon-based sentimental analysis assigns sentimental scores or labels to each token in a predefined library, meaning that it cannot classify words that are not in the library. However, sentimental lexicons are usually constructed with manual annotation on carefully chosen tokens by linguists, so these lexicons are usually more reliable for sentimental tasks when there is insufficient training data for the machine learning models. Specifically, we used the sentimental lexicon NRC created by Saif Mohammad and Peter Turney [36, 37]. The NRC lexicon assigns various sentimental labels to each word, and it includes some of the most frequently used English nouns, verbs, adjectives, and adverbs. Of course, the NRC lexicon does not include most of the Covid-related terms, so we used the bigram data to find out the in-bag tokens associated with Covid-related terms to assign a sentimental score for them. The details of the previous scheme can be found in Appendix A.

#### 3.2.3 Statistical model selection and validation

The purpose of using machine learning is to select the most important variables and then build a parsimonious and predictive model from these variables to predict social media fluctuation. As mentioned earlier, we used the year 2021’s data for model selection and calibration and the year 2022’s data for validation. We used a nested cross-validation approach on the year 2021’s data to perform feature and model selections. As shown in Figure 3, we divided the year 2021’s data such that there are five outer folds and five inner folds for each outer fold. To reduce the search space, we used the inner folds to select the features and the outer folds to select between models with different feature encodings and transformations. Features and models are selected based on the test MSE and one-standard-error rule to obtain robust selection results that are both parsimonious and predictive. Finally, we tested the justified model on the year 2022’s data.

**Figure 3:**
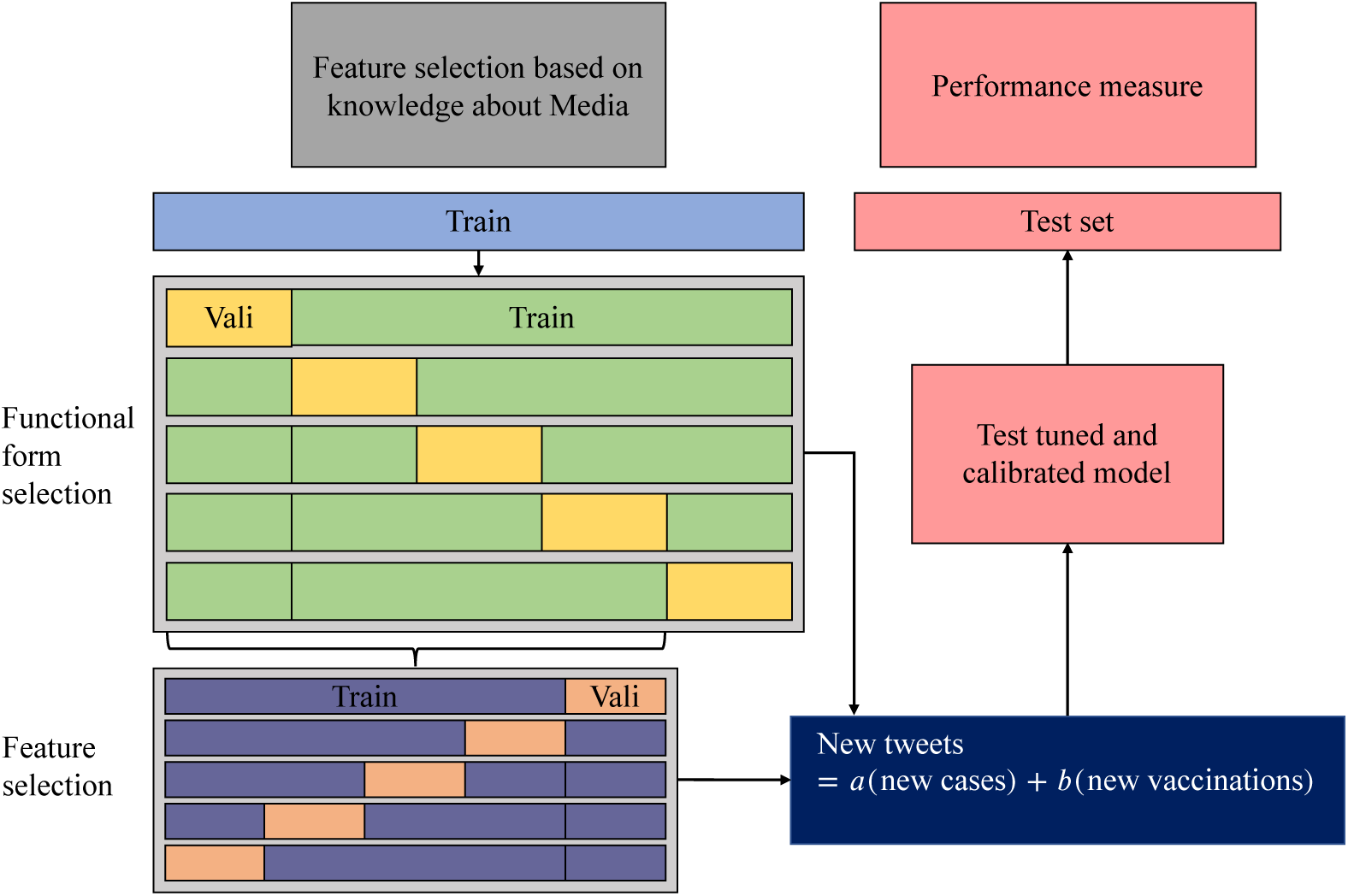
Nested cross-validation scheme on 2021’s data for feature and functional form selections. We used the 25 inner folds to select the most important features to predict social media fluctuation, and we then used the 5 outer folds to select the most predictive functional form for these important features. Once we were done with the model selection, we calibrated the final differential equation model on the entire 2021’s data. The test set is 2022’s data, and we tested our final differential equation model on this set.

#### 3.2.4 Bayesian inference

Our inferential process consists of three major steps: fitting, testing, and prediction. In addition, We considered two nondisjoint and inter-dependent time-varying factors as our prior knowledge during the full course of the process: vaccine availability and disease variant. In the fitting step, we fitted our differential equation model (equation 1) to the entire 2021 data together with a part of the 2022 data to infer the values of time-homogeneous parameters and time-dependent parameters. Specifically, we chose parameter *γ*, *δ*, *r*, and *v_C_* to be time-dependent, where the first three parameters are mainly controlled by the properties of the currently dominant disease variant, and the last one is affected by the development and production of vaccine in response to different disease variants. We considered the prevalence of the Omicron variant as the turning point for all four time-dependent parameters, where they change from one set of constant values ({*γ, δ, r, v_C_*}) to another set of constant values 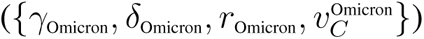. In addition, we reset the recovered pools to the corresponding susceptible pools at the point (Figure 2) when the Omicron variant becomes prevalent.

We used our model to compute the expected weekly incidence, the expected weekly number of vaccinations, and the expected weekly number of tweets. Then, we used negative binomial distribution as the likelihood model to account for the potentially large observational errors in the data (details can be found in Appendix C). We used *Stan*, a programming language for Hamiltonian Monte Carlo (HMC) Bayesian inference, to obtain the posterior distributions and credible intervals of the parameters and initial conditions (https://mc-stan.org).

For the testing step, we obtained the posterior predictive distributions from the calibrated and compared the model prediction with the rest of the 2022 data that is not used for model calibration. Finally, we used the calibrated model to predict the future dynamics and explore some intervention strategies to examine their impact on disease transmission.

## 4 Result

### 4.1 Data analysis

The detailed results of text mining and sentimental analysis can be found in Appendix A. In short, we demonstrated that Covid cases, vaccines, and deaths are overrepresented in social media content, indicating that these factors are potential drivers of disease-related social media (Figure S1). In addition, we showed that the proportion of appropriate media stays roughly constant throughout time, and the proportion of inappropriate rigid media is negligible throughout time (Figure S2).

The detailed results of feature and model selection can be found in Appendix B. In sum, we found that the following linear equation carries the most predictive power for the new media, so we integrated this equation into our final model (Equation 1), to predict the rate of change of disease-related media 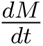.

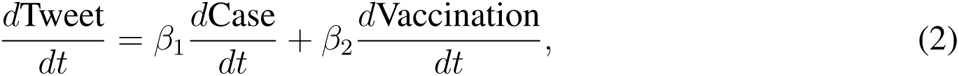

In plain words, the newly generated social media is driven by the new infections and new vaccinations.

### 4.2 Sensitivity analysis

Our model contains 7 variables and 9 parameters, and all the variables are not directly observable, so our first step is the sensitivity analysis, which can help us identify the most significant inputs, some can be used for the preparedness of the pandemic.

We evaluated the model sensitivity with the metric named partial rank correlation coefficient (PRCC), which measures the monotonicity between each of the parameters and the user-specified response. Following the well-accepted procedure for PRCC, Latin hypercube sampling and partial ranked correlation were used to evenly sample in the feature space and to prevent confounding effects that lead to false positive associations. We selected two response metrics: cumulative number of cases and cumulative number of vaccinations. A positive or a negative PRCC indicates a positive or negative monotonic relationship between the response and one parameter after controlling all the other parameters. The more positive or negative the PRCC is, the stronger the monotonic relationship is.

As shown in Figure 4, the cumulative number of cases is sensitive to several behavior-related parameters, such as the maximum behavioral transition rate (A), reduced transmission ‘compliant/non-compliant’ (*ω*), compliant vaccination rate (vC), and threshold for noncompliance (MT). Specifically, the increase in the behavioral transition rate (A) will lead to lower cumulative incidence, and the increase of the reduced transmission ‘compliant/non-compliant’ (*ω*), meaning that the compliant hosts take fewer effective preventive measures, will cause higher cumulative incidence. A larger MT causes a faster loss of compliance and therefore will lead to a higher cumulative incidence. One may expect an increase in the compliant vaccination rate (vC) can effectively reduce the cumulative incidence, but vC only has a moderate effect because it initiates the negative feedback loop: the more compliant hosts, the more hosts get vaccinated, which leads to faster loss of compliance and eventually slows vaccination down. In addition, media-relative parameters such as infection-driven media change (*ɛ*) and vaccination-driven media change (*σ*) are negatively associated with the cumulative incidence, suggesting that media can potentially be a disease control mechanism. Moreover, the initial numbers of noncompliant susceptibles and noncompliant infectives are very sensitive, suggesting that increasing hosts’ compliance prior to the pandemic may effectively reduce the size of the pandemic.

**Figure 4:**
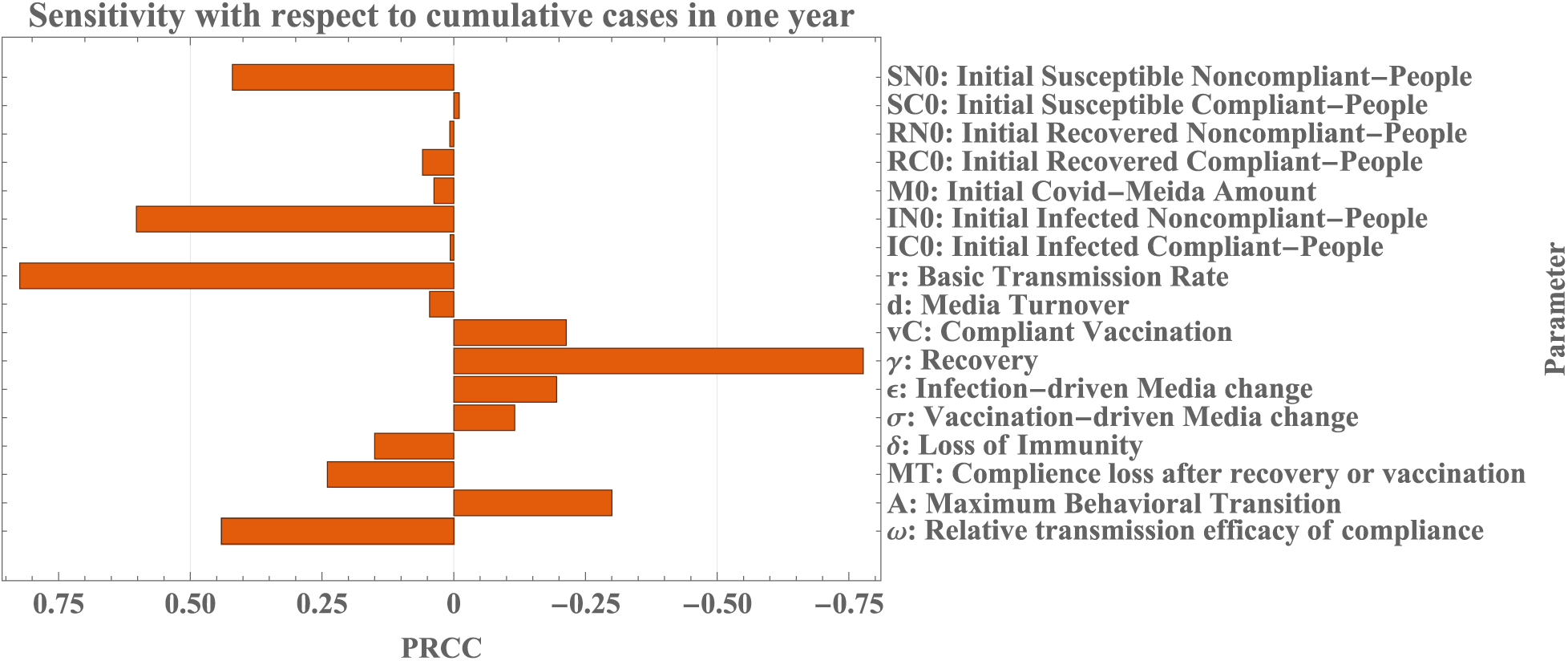
Sensitivity according to the cumulative cases in one year. The higher the PRCC value, the strong the monotonicity between the parameter and the cumulative cases.

Sensitivity analysis with respect to the cumulative number of vaccinations shows somewhat consistent but slightly different results. As shown in Figure 5, the cumulative number of vaccinations is relatively less sensitive to A and *ω*, but it remains sensitive to MT as the loss of compliance can reduce the number of vaccinations. Moreover, the cumulative number of vaccinations is very sensitive to the vaccination-driven media change (*σ*) and media turnover (d), indicating that media is important in maintaining compliance among the population and thereby the vaccination.

**Figure 5:**
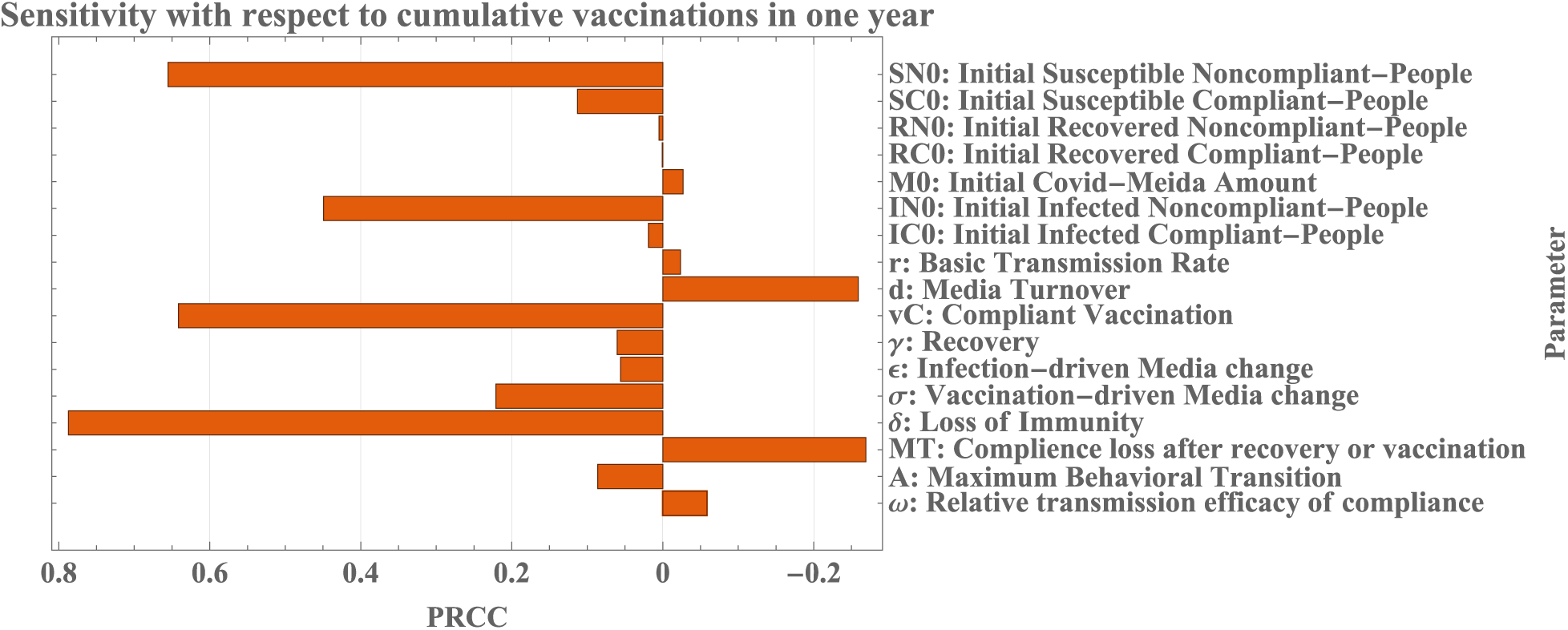
Sensitivity according to the cumulative vaccinations in one year. The higher the PRCC value, the strong the monotonicity between the parameter and the cumulative vaccinations.

### 4.3 Model calibration and validation

To further understand the actual effects of both behavior and social media inputs, we fitted and validated our model to the data according to the scheme in section 3.2.4. Our BMSIR model offers a much better fit and a more accurate prediction than the standard SIR model. Although we didn’t directly validate the behavior components in our system, our simple model produces reasonable predictions of incidence, vaccination, and social media, simultaneously. However, our simple model fails to capture all the aspects of the data, which is reasonable as human interventions were As shown in Figure 6, for the weekly data in the year 2021 the model prediction roughly recovers the pattern in the incidence data, but the model only captures part of the patterns in the vaccination and media data. Specifically, the major discrepancies between the model-predicted incidence and the data occur between January and July, and the model fails to capture two sudden drops in cases and one sudden increase in vaccination. These discrepancies can be attributed to the governmental interventions during that period, such as mandatory vaccinations, increased testing and surveillance, travel policies, quarantine, and mandatory preventative measures in workplaces.

**Figure 6:**
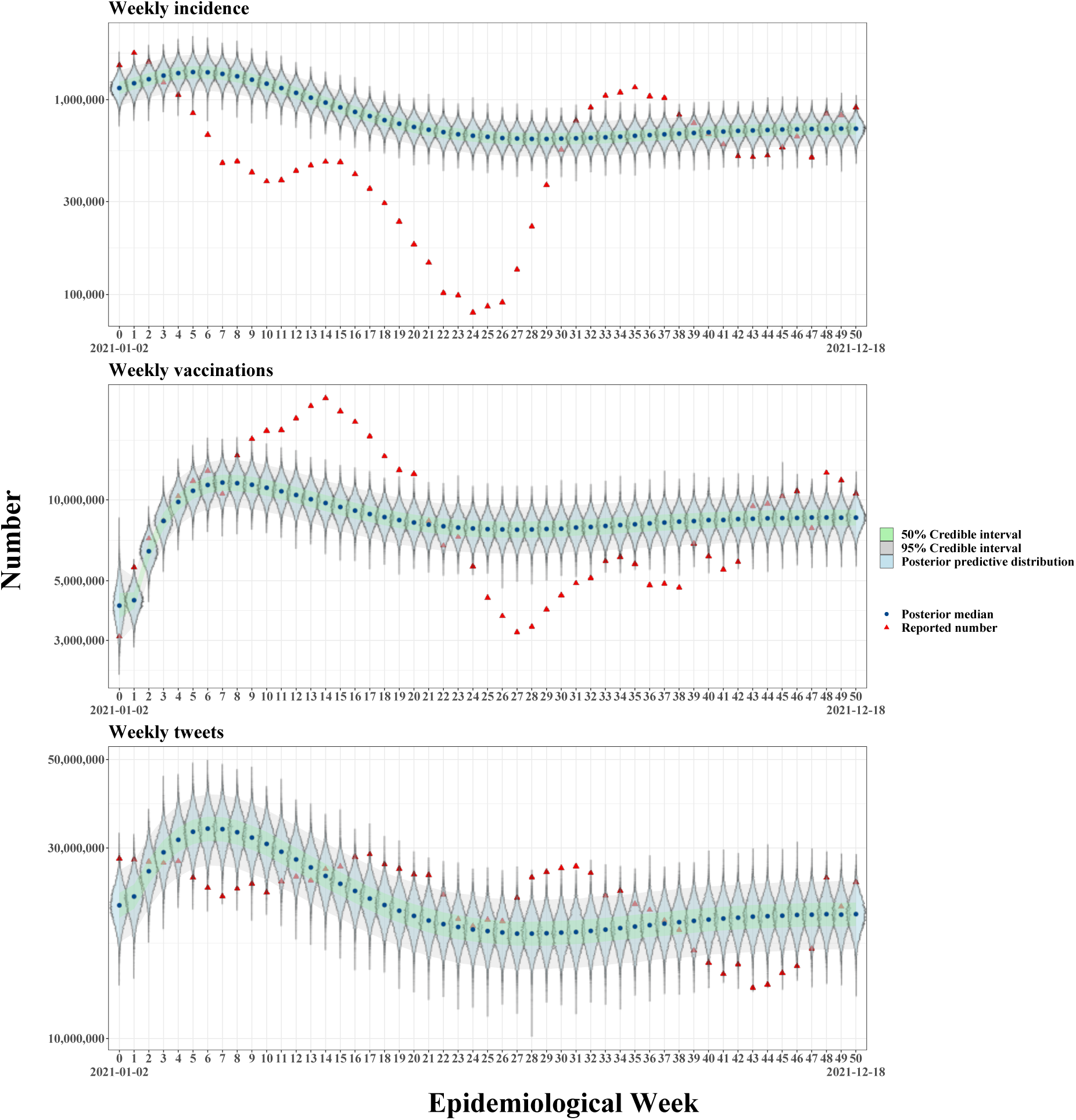
Bayesian posterior predictive distributions for the weekly data in the year 2021 using the BMSIR model. We used all the data points in this figure for the Bayesian inference.

Again, for the weekly data in the year 2021 and year 2022, the model captures most of the pattern in the incidence data, and the model predictions for media and vaccination roughly follow the trends in the data. The inaccurate predictions are possibly due to the drastic difference between Omicron and the variants in 2021. Also, the current model assumes sufficient vaccine supply at all times, but the Omicron-specific vaccine was released at the end of August 2022.

The detailed posterior parameter estimates for the BMSIR model can be found in Appendix C. Figure S6 shows the posterior distributions of the parameters in our model. The posterior distribution of *ω* shows that compliant individual, on average, has roughly a 20.0%-22.4% reduction in their exposure and transmissibility. The duration of immunity 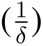 for alpha and delta variants is about 74.7-84.7 days, approximately 2 and a half months. For Omicron, the duration of immunity is about 147-168 days, slightly less than half of a year. Thus, immunity to the Omicron variant stays approximately two times longer compared to the immunity to the Beta and Alpha variants. By comparing the posterior statistics of *σ* and *ɛ*, we can see that one new vaccination leads to a significantly higher production of disease-related media compared to one new incidence (about 14 times higher). The posterior infectious/recovery period 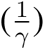 for alpha and delta variants is about 8.25-8.65 days, and the infectious/recovery period for the omicron variant is about 4.13-4.34 days. Thus, the average infectious period for Omicron is shorter than those of Alpha and Beta variants. The posterior distribution for *r* indicates that the Omicron variant is approximately 1.65 times more transmissible than the Beta and Alpha variants. In addition, posterior inference on parameter *vC* shows that the rate of taking the booster vaccine is lower than the rate of taking the regular vaccine, which is consistent with the delayed release of the Omicron-specific vaccine. Finally, the posterior distribution for *d* shows that disease-related media remains effective on the social media platform, on average, for 21-24 days. Our estimated infectious period and immunity duration of Alpha and Beta variants are mostly consistent with the estimates (9 days for the infectious period and 0.5-10 years for the immunity duration of Alpha and Beta variants) by Lavine et al. (2023) [38]. Figure S7 shows the posterior distributions of the initial conditions. Our fitting result indicates that at the beginning of 2021, there are roughly 70.0-72.0%, 0.381-0.510%, and 27.6-29.5% susceptible, infected, and recovered hosts. The initial proportions of compliant hosts in the S, I, and R pools are about 4.86-11.5%, 4.12-98.0%, and 3.62-97.2%, which are quite diffusive, possibly due to the small sample size for training.

To justify the important roles of behavior and media in the disease transmission process, we compared our behavioral-modified SIR (BMSIR) model with the standard SIR model, whose fitting results are in Appendix C. By comparing Figure 6 and S8, one can see that the BMSIR model has a better agreement with the data compared to the standard SIR model, suggesting that the BMSIR model is more flexible and presumably less biased compared to the standard SIR model. Granted, one might argue that the better fit of the BMSIR model is due to its higher degrees of freedom compared to the standard SIR, but we wanted to show that this conclusion holds also in the test data part. By comparing Figure 7 and S9, we can see that the standard SIR model fails to capture the endemicity and overestimates the weekly number of vaccinations, and the BMSIR model can still enclose most of the test data in its credible intervals. In sum, the BMSIR model seems to be much more similar to the actual data generation process compared to the standard SIR model, indicating that behavior and media are likely to play important roles in the disease transmission process.

**Figure 7:**
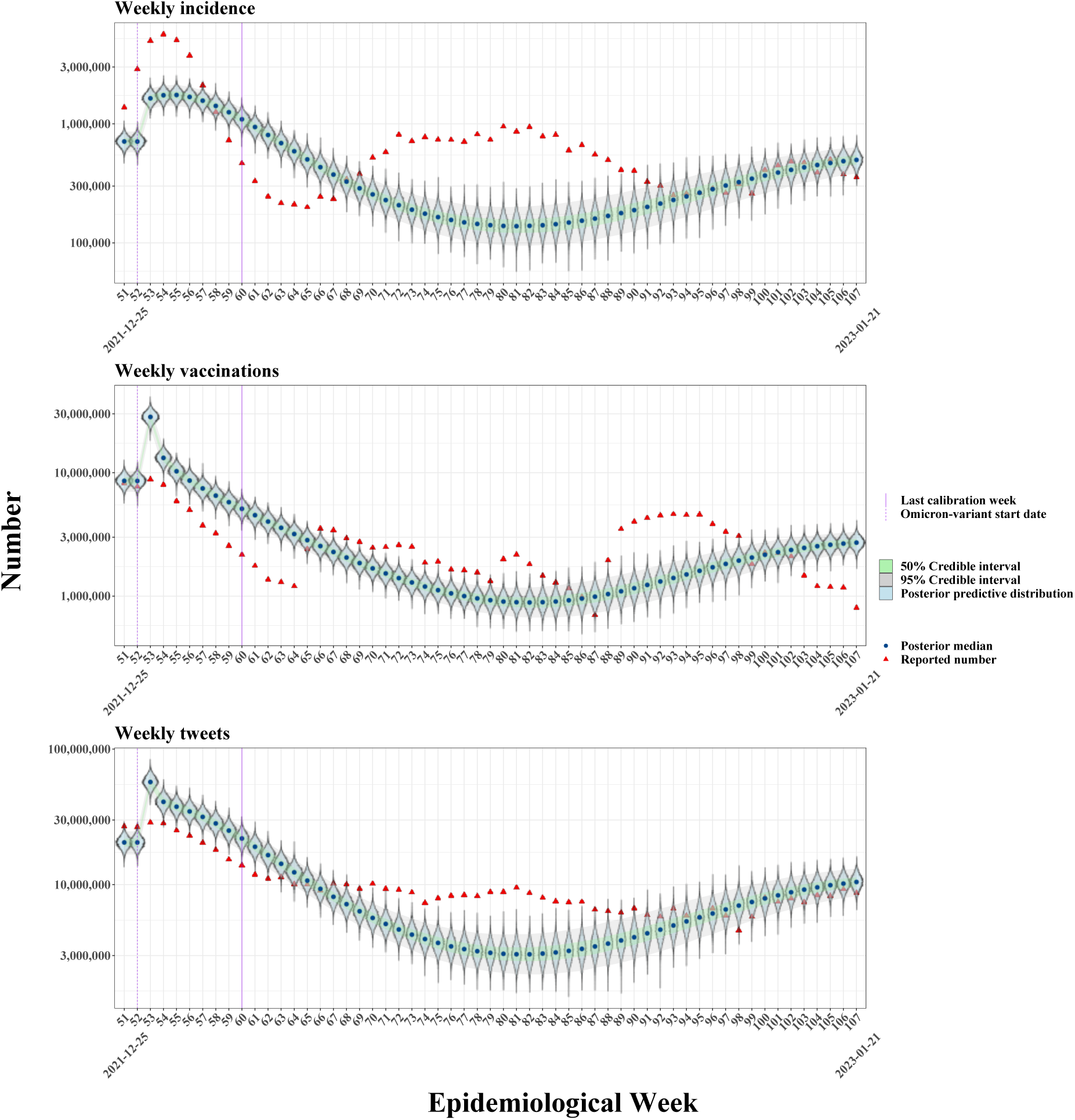
Bayesian posterior predictive distributions for the weekly data in the year 2021 and year 2022 using the BMSIR model. In week 52, we moved subpopulations in the recovered pools to the corresponding susceptible pools, and some model parameters start to use Omicron-specific values. Week 60 is the last data point used for training, and the model is fully blind to all the data points after week 60. The data after week 60 is the test part, and we simulated the model trained on data before and include week 60 to compute the posterior predictive distributions of the data after week 60.

### 4.4 Prediction and control

#### 4.4.1 Effects of behavioral and media inputs on the pandemic outcome

Using the posterior knowledge from section 4.3, we explored the effects of different behavioral and media inputs on the US pandemic outcomes. The first parameter of interest is the maximum behavioral switching rate (A), which reflects hosts’ average flexibility in their behaviors. As shown in Figure 8, when A is nonzero, the more behaviorally flexible the hosts are, the less incidence and more vaccination there are in approximately one year. In addition, the calibrated model shows that increasing behavioral flexibility can reduce the yearly prevalence of infection to approximately 12% of the total US population and increase the prevalence of vaccination to approximately 1.3 per person. Interestingly, when hosts are maximally rigid (A = 0), the yearly prevalences of incidence and vaccination will both be low.

**Figure 8:**
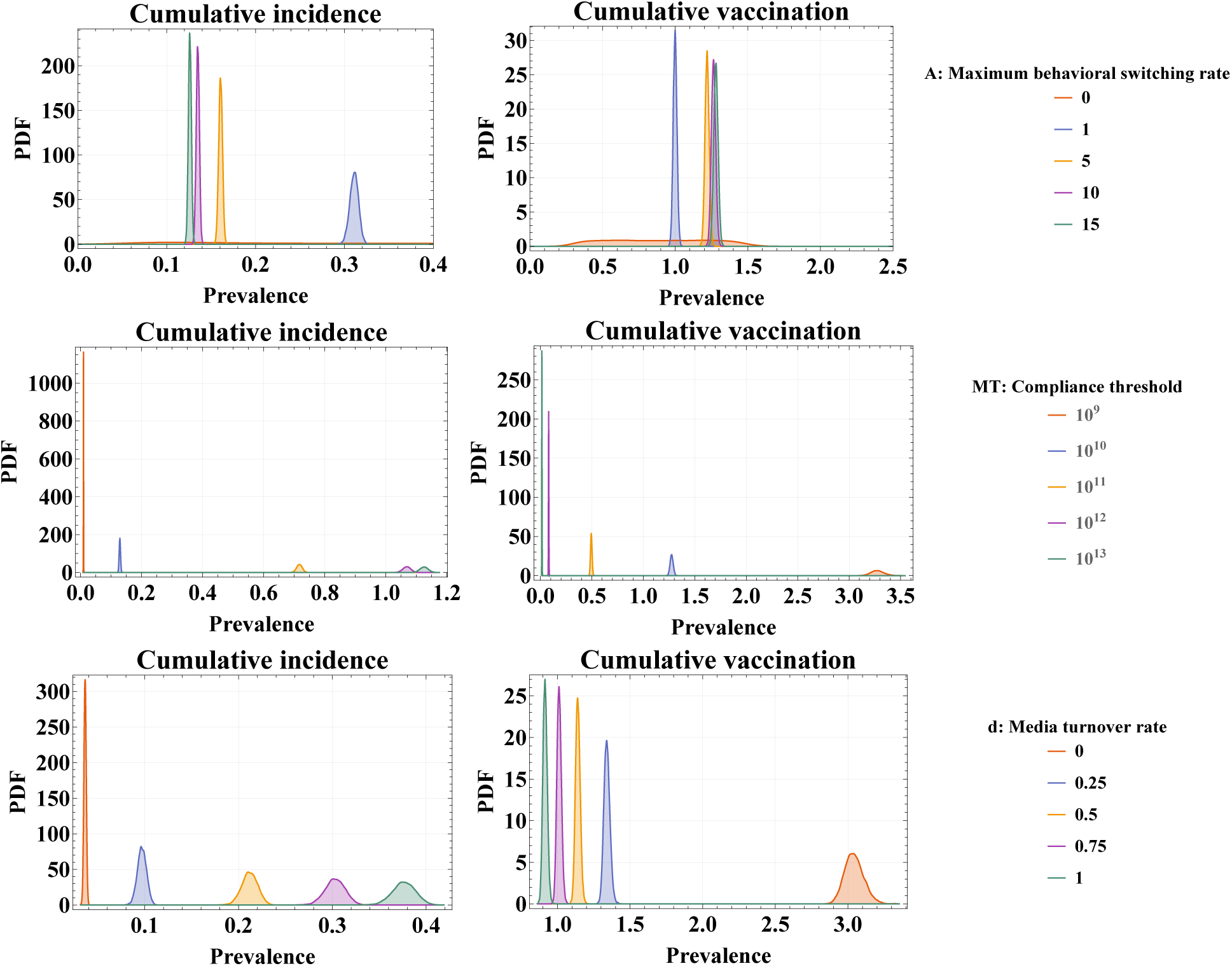
Distritbuions of the yearly cumulative incidence and yearly cumulative vaccination using posterior knowledge of the BMSIR model from section 4.3 after altering the value of parameter *A*, MT, and *d*.

Next, we altered the value of the compliance threshold (MT), which reflects hosts’ sensitivity to media. When hosts become less sensitive to media (larger MT), the incidence will increase, and the vaccination will decrease. As shown in Figure 8, when hosts’ sensitivity to media drops to a certain level, hosts will almost stop taking vaccination, and the yearly prevalence of infection will grow above 100%. This is a realistic problem as hosts’ distrust tends to build up during the pandemic, and our result suggests that such distrust can be detrimental.

Lastly, we altered the value of the media turnover rate, which reflects the average lifespan of the disease-related media. As shown in Figure 8, the shorter the life span of the disease-related media (larger d), the more incidence, and less vaccination there will be. Our result shows that increasing the lifespan of the disease-related media can substantially reduce the size of the outbreak.

#### 4.4.2 Control strategy

To further explore the effects of behavior and social media on some hypothetical disease control strategies, we computed and extended the posterior trajectories of the variables in our BMSIR model until the year 2025 by using the posterior knowledge from section 4.3. Figure 9 shows the posterior trajectories of the model variables and weekly variables derived from model variables without any additional intervention strategies. The second column of Figure 9 basically repeats the information in Figure 6 and 7 but shows instead the posterior distribution of the expected weekly incidence, weekly vaccinations, and weekly tweets, while Figure 6 and 7 shows the posterior predictive distribution of the actual observed data. In Figure 9, trajectories continue until October 2025, and we assume no new variants that are substantially different from Omicron. Our model predicts that the proportions of compliant hosts in different epidemiological compartments remain transiently high only at the beginning of the outbreak and keep constantly low for the rest of the time. Also, our model predicts that Covid-19 will become endemic with approximately 0.1% of the total US population if there are no additional perturbations, such as better treatments, surveillance, or new variants. Starting from this simulation, we explored two hypothetical control strategies: better preventative measures and governmental awareness programs.

**Figure 9:**
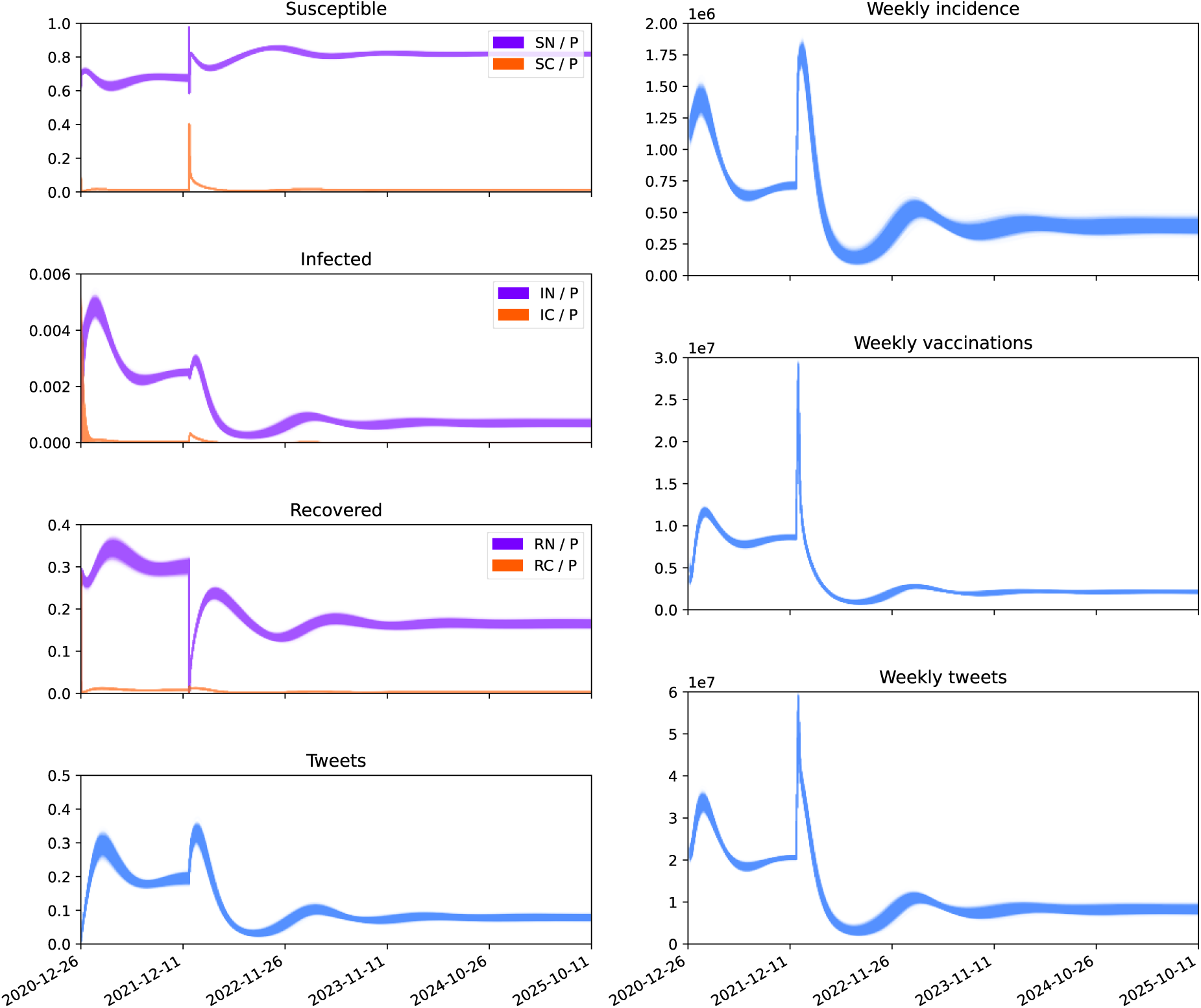
Posterior trajectories of the BMSIR model from section 4.3. In the left column, we showed the proportions of both behavioral pools of susceptible, infected, and recovered hosts in the total population. For the instantaneous tweets, we showed the number of tweets per person. In the right column, we showed weekly incidence, vaccinations, and tweets in absolute numbers.

We first altered parameter *ω* as an analogy to having better preventative measures while keeping other things intact. In the new simulation, we reduced *ω* by half starting from January 20, 2024 (160 weeks from December 26, 2020), meaning that compliant hosts will have more and better choices of preventative measures to reduce their exposure and transmission. As shown in Figure 10, having more and better preventative measures does not significantly reduce the infection, and our model simulation suggests that this is due to the extremely low proportion of compliant hosts in the population after the pandemic.

**Figure 10:**
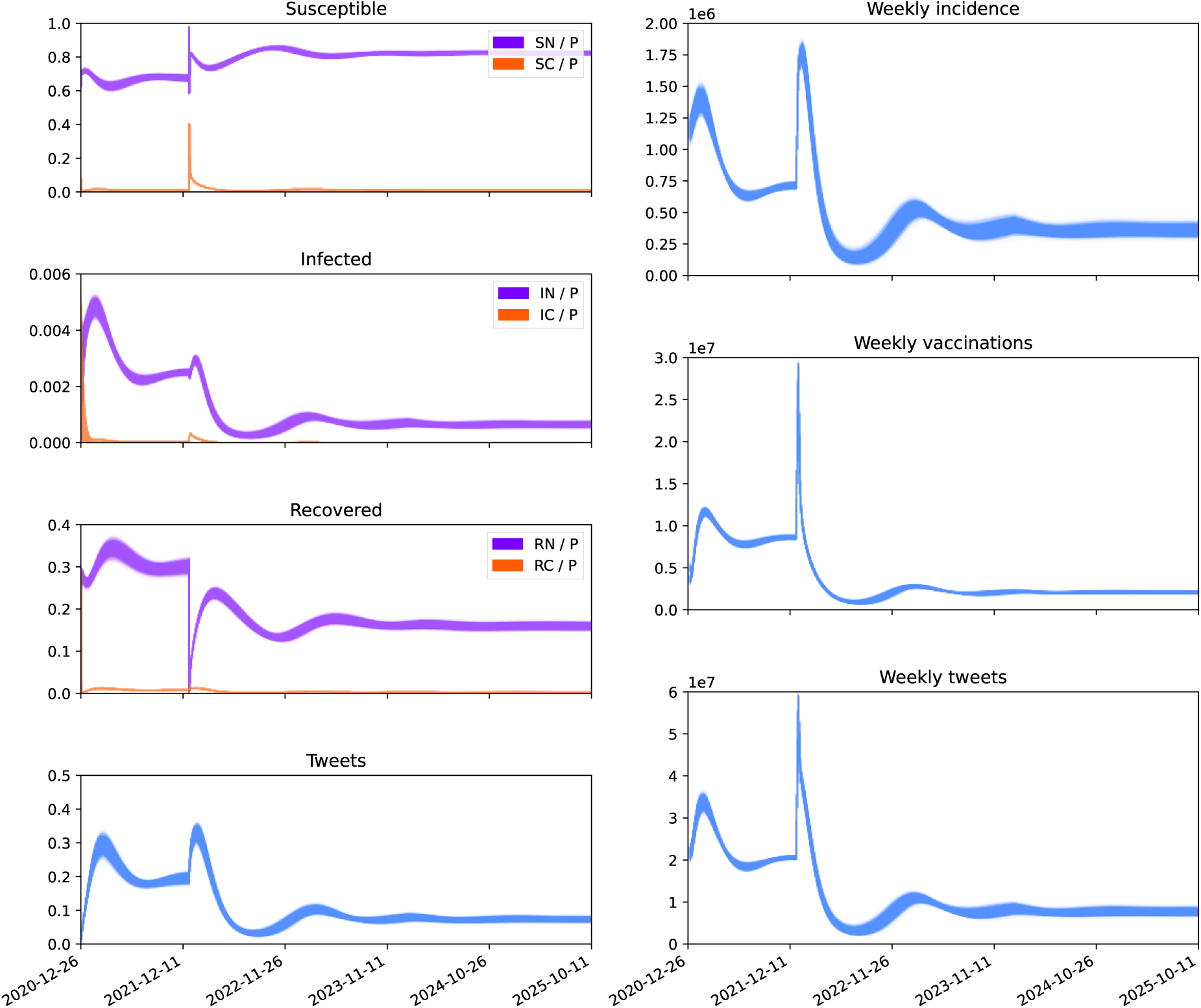
Posterior trajectories of the BMSIR model from section 4.3 but with addition and better preventative measure since January 20, 2024 (160 weeks from December 26, 2020). Specifically, parameter *ω* decreases by half starting from week 160.

Next, we added an additional sourcing term *g*(*t*) to 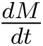 to represent the information about the awareness programs on social media platforms. Specifically, we let the awareness programs for infectious disease to happen every four weeks and continue to exist for one week, starting from January 20, 2024 (160 weeks from December 26, 2020). We therefore modeled *g*(*t*) as:

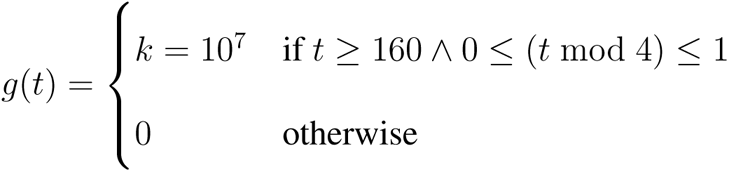

If such a control scheme exists, there will be 70 million additional awareness-program-related tweets in each four-week period, which is really a minuscule amount considering that there are roughly 500 million new tweets per day (these additional awareness-program-related tweets only take about 0.5% of all the new tweets in four weeks). Surprisingly, such minor media input is much more effective in disease control than our previous control scheme that reduces compliant exposure and transmissibility by half. As shown in Figure 11, additional media input is more effective than the additional preventative measures in reducing the infection and endemicity, and it also increases the number of vaccinations. Our model simulation indicates that, after the pandemic, interventions through the media are more effective than direct preventative measures, and preventative measures will be more effective when there is a sufficient amount of compliant hosts in the whole population.

**Figure 11:**
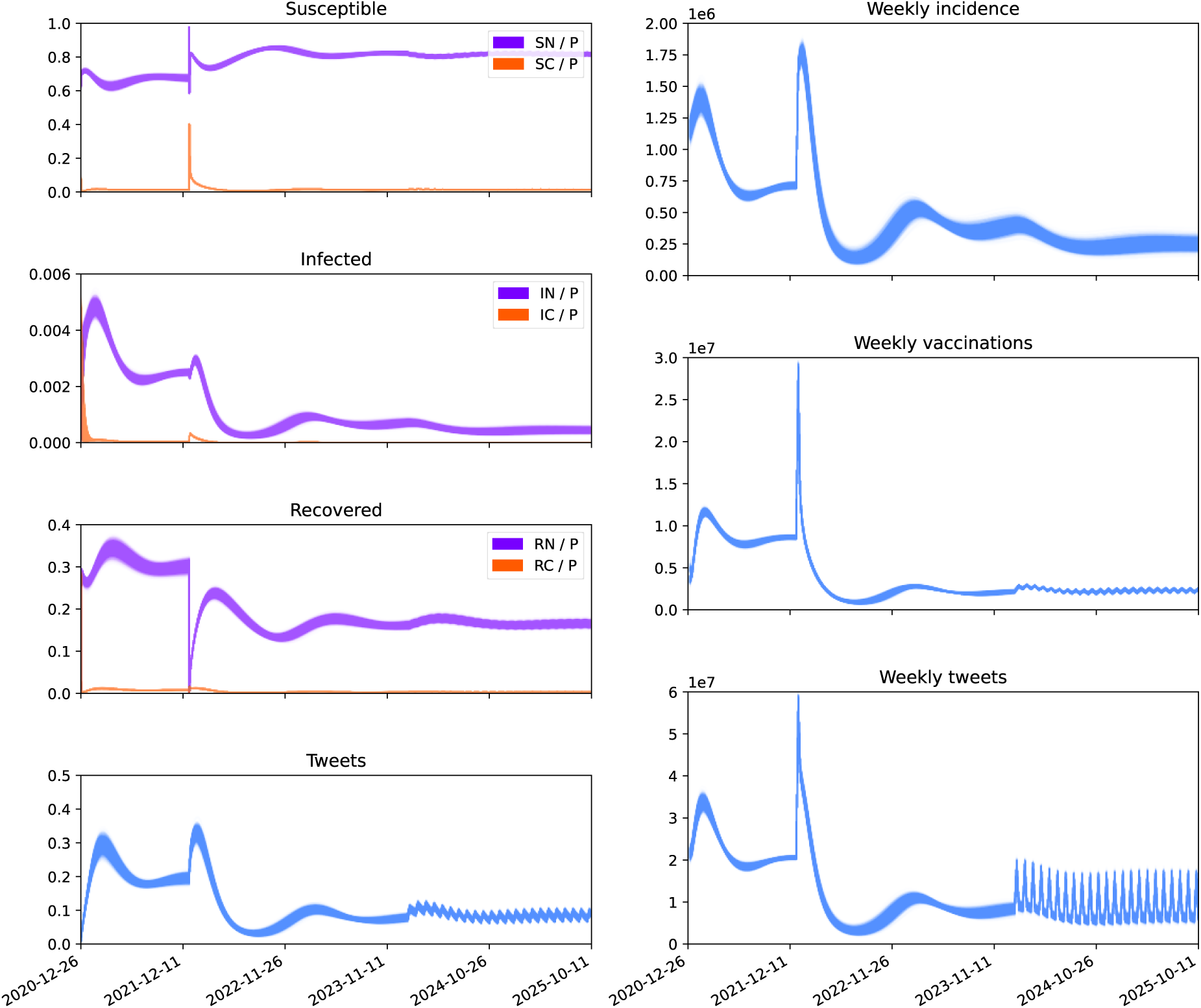
Posterior trajectories of the BMSIR model from section 4.3 but with information about awareness program as additional input to social media. Specifically, we added an additional term *g*(*t*) to 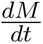 starting from week 160. We modeled *g*(*t*) such that the input to media activates every four weeks and keeps active for one week after each activation.

## 5 Conclusion and discussion

Human behavior and social media are two important factors in determining the pandemic outcomes, and many researchers have incorporated human behavior into their predictive models [14–23, 25–28, 39]. However, there is currently no gold standard to model the interplay between human behavior, infection, and social media. Many previous works have provided theoretical frameworks to model the behavioral and social media inputs, but these studies lack data validation to justify their assumptions.

In this study, we combined data analytic techniques and mathematical modeling in order to address three major questions. First, how to incorporate behavior and social media into the standard SIR model? Although many past works have demonstrated approaches to model human behavior, the focus on social media is insufficient. By using a nested cross-validation scheme and feature and model selection techniques, we demonstrated that the most efficient form to predict the weekly new disease-related tweets is the linear form with two main effects: weekly new incidence and weekly new vaccinations. We, therefore, incorporated media into the standard SIR model according to this linear form. For the behavior part, we borrowed some ideas from Funk et al. (2010) and Misra et al. (2011) with some modifications [24, 25]. Then, we used many approaches such as sentimental analysis, data calibration, data validation, and model comparison to justify some of our assumptions and part of the model formulation. Indeed, our model is still rather simple and cannot capture all the patterns in the data, and we didn’t validate some aspects of the model, such as the noncompliance and compliance separation. Unfortunately, validating human behavior is a challenging task as human behavior is not directly observable, but with the growing dimensionality of the data, one can still try to infer some of the behavior variables. In this study, we used the overall model test accuracy to justify our framework, but the correctness of the model still requires validation on each individual part.

One of our main questions is whether behavior and media play significant roles in the transmission of the disease. Through the data validation, we have shown that incorporating behavior and media significantly improves the fit on both the training and testing part compared to the standard SIR model. In addition, our sensitivity analysis indicates that infection and vaccination are very sensitive to behavior and media inputs.

Another question is how behavior and media affect the pandemic outcomes. With our posterior knowledge derived from the US Covid-19 data, we manipulated different parameters to demonstrate the effects of behavior and media on a somewhat realistic setting. We demonstrated that behavioral flexibility, media sensitivity, and media lifespan can significantly affect infection and vaccination. Moreover, we showed that due to the limited proportion of compliant hosts in the population, having more and better preventative measures is not as effective as having more awareness programs, as the latter promotes hosts’ compliance to better reduce the infection and increase the vaccination.

Future investigations can explore media stratification, differential human attention, and behavioral rigidity. In addition, future studies should focus more on how to obtain relevant data to validate human behavior.

## Data Availability

All data produced in the present study are available upon reasonable request to the authors

## 6 Acknowledgment

D.G. was supported by NSF (FAIN) 2200255 grant.

## Appendix

## Appendix A: Twitter analysis

As shown in Table S1, we investigated six general Covid topics and assigned some topic-specific terms to them. Next, for each topic-specific term, we filtered out the associated in-bag tokens from the bigram data. In addition, we assigned the appropriate, inappropriate nonrigid, and inappropriate rigid sentiments to each topic, and we investigated the composition of these three categories for each topic-specific term and eventually the topic. Here, “appropriate” means that the media can promote hosts’ compliance, and “inappropriate” means that the media encourages non-compliance. Moreover, “rigid” means that media can increase hosts’ distrust and insensitivity to the appropriate media and therefore increase the inertia of non-compliant hosts. Since social media always vary with time, we picked four months (January, April, July, and November) in the year 2021 and 2022, and we repeated the analysis on each of these time points. To give an example, suppose “vaccine” is associated with “effective”, then the proportion of appropriate sentiment is 1. If “vaccine” is associated with “inequality” the proportion of inappropriate nonrigid sentiment is 1. If “vaccine” is associated with “injury” the proportion of inappropriate nonrigid sentiment is 75%, and the proportion of inappropriate rigid sentiment is 25%.

**Table S1:**
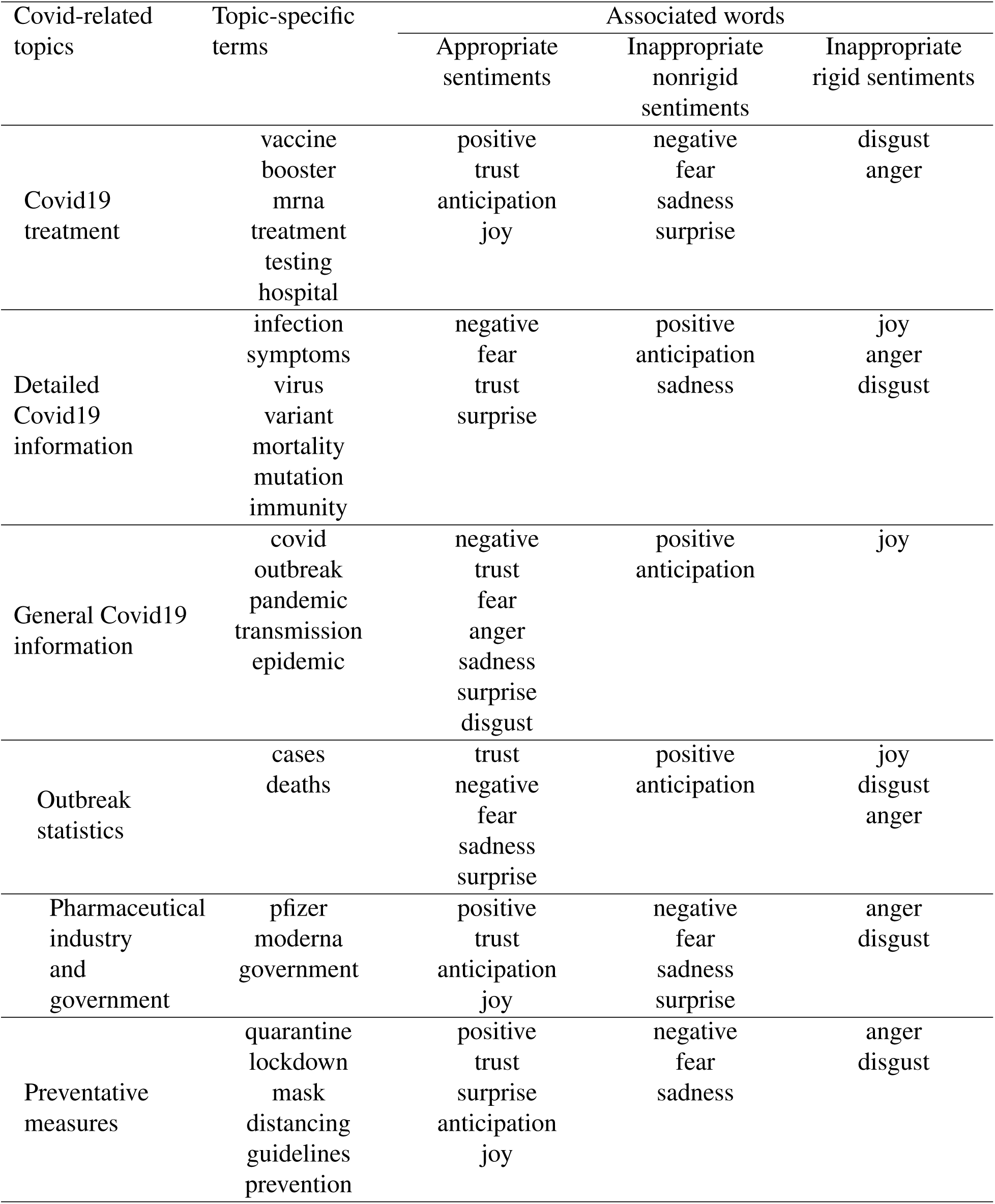
Sentimental analysis of the content of the Covid-related tweets. We investigated six general Covid topics by picking their terms. For each topic-specific term, we obtained its associated words and categorized their sentiments into three classes in terms of the current context. Each word can have mixed sentiments, so we computed the expected amounts of appropriate, inappropriate nonrigid, and inappropriate rigid sentiments for each word, each topic-specific term, and finally each Covid-related topic.

As mentioned earlier, we assumed that the quantity of Covid-related social media and the expected recovery state affect the gain and loss of compliance, and the pandemic state drives the change in the quantity of Covid-related social media. To justify this assumption and discover possible driving factors of Covid-related social media, we did text mining on the Twitter data. As shown in Figure S1, the mostly-used terms in the Covid-related tweets indicate that in 2021, Covid cases, vaccines, and deaths are the three highly popular topics besides Covid itself. This suggests that content creators on social media were more interested in these topics and produced more tweets related to these topics, leading to the overrepresentation of these terms in the tweet content. Thus, cases, vaccines, and deaths are three possible drivers of social media. In 2022, although the previous three topics were still among hosts’ major focus, some new trends appeared in the content. For instance, the prevalence of the Omicron variant around the start of 2022 has gained a lot of attention on social media for more than half of a year, coupled with the increasing popularity of the booster vaccine. Granted, in order to accurately predict the dynamics of social media, one has to account for the emergence of such “breaking events,” like Omircon. However, incorporating random events into the predictive model requires sufficient biological understanding and support, which is usually impossible to obtain at the beginning of the pandemic, and such a specifically-tunned model may have poor generalizability.

**Figure S1:**
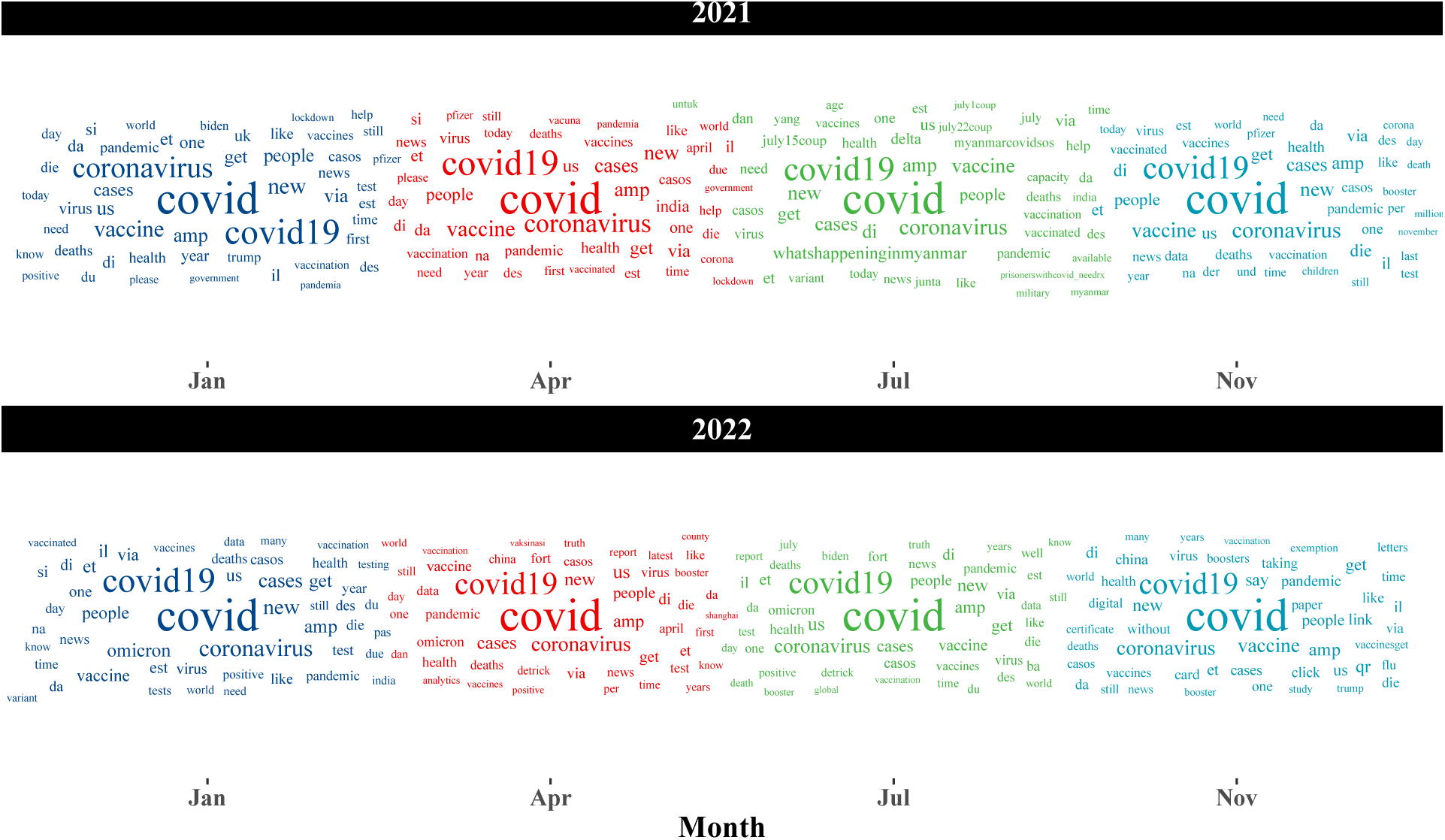
Wordclouds for the top unigrams from the Covid-related tweets in 2021 and 2022

Next, to ensure that using the media quantity and excluding the media content is a valid assumption, we did a lexicon-based sentimental analysis using the NRC emotion lexicon. One limitation of the lexicon-based sentimental analysis is its inability of assigning sentiments to the out-of-dictionary words, and in our case, most Covid-related terms are indeed not included in the NRC dictionary. Thus, we focused on the top bigrams and found the within-dictionary words associated with the Covid-related terms, and we calculated the proportions of sentimental appropriateness and inappropriateness among these associations as approximations to the proportions of appropriate (compliance-boosting) and inappropriate (compliance-inhibiting) media (note that one tweet can have mixed content), for six different Covid-related topics and their union across four months in both 2021 and 2022. In addition, we calculated the proportion of sentimental rigidity for every bigram association, representing individuals and media outlets that strongly oppose the appropriate media. We expect the proportion of inappropriate rigid media to be as small as possible so that the majority of the population could still respond actively to Covid-related media during the pandemic.

As shown in Figure S2 A, in 2021, we got over 60% appropriate media in all four months, and the appropriateness of the media slightly decreases in January, April, and July but slightly increased in November of 2022. Also, the proportion of inappropriate rigid media is negligible at all the sampling points. Thus, the proportion of appropriate media roughly stays constant during the pandemic, so we assumed that the overall quantity of all Covid-related media itself is sufficient to predict the behavioral change, as the proportion of appropriate media is roughly a constant scaling factor throughout the pandemic. As for the inappropriate rigid content, since it only takes a relatively small proportion, we assumed that the amount of behaviorally rigid hosts is negligible, and their tweets are too few to affect other hosts’ thoughts and behavior.

In Figure S2 B, we summarized the proportions of our media categories in each Covid-related topic. In 2021, Covid treatment seems to have the most appropriate content compared to the other topics, and the outbreak statistics (e.g. cases and deaths) have the least appropriate content during the middle of the year, suggesting that individuals and media were relatively more positive and optimistic on the Covid treatment (e.g. vaccine and drug) while being relatively more negative and distrust on the outbreak statistics. In 2022, the appropriate content for Covid19 measures is relatively lower compared to that of 2021, possibly due to the fast emergence of different Covid variants, especially the Omicron variant that can escape the previously established immune surveillance. Meanwhile, the pharmaceutical industry and government in 2022 have the most appropriate content, suggesting an increased trust and hope for novel industrial products (e.g. booster vaccine) and better governmental efforts.

**Figure S2:**
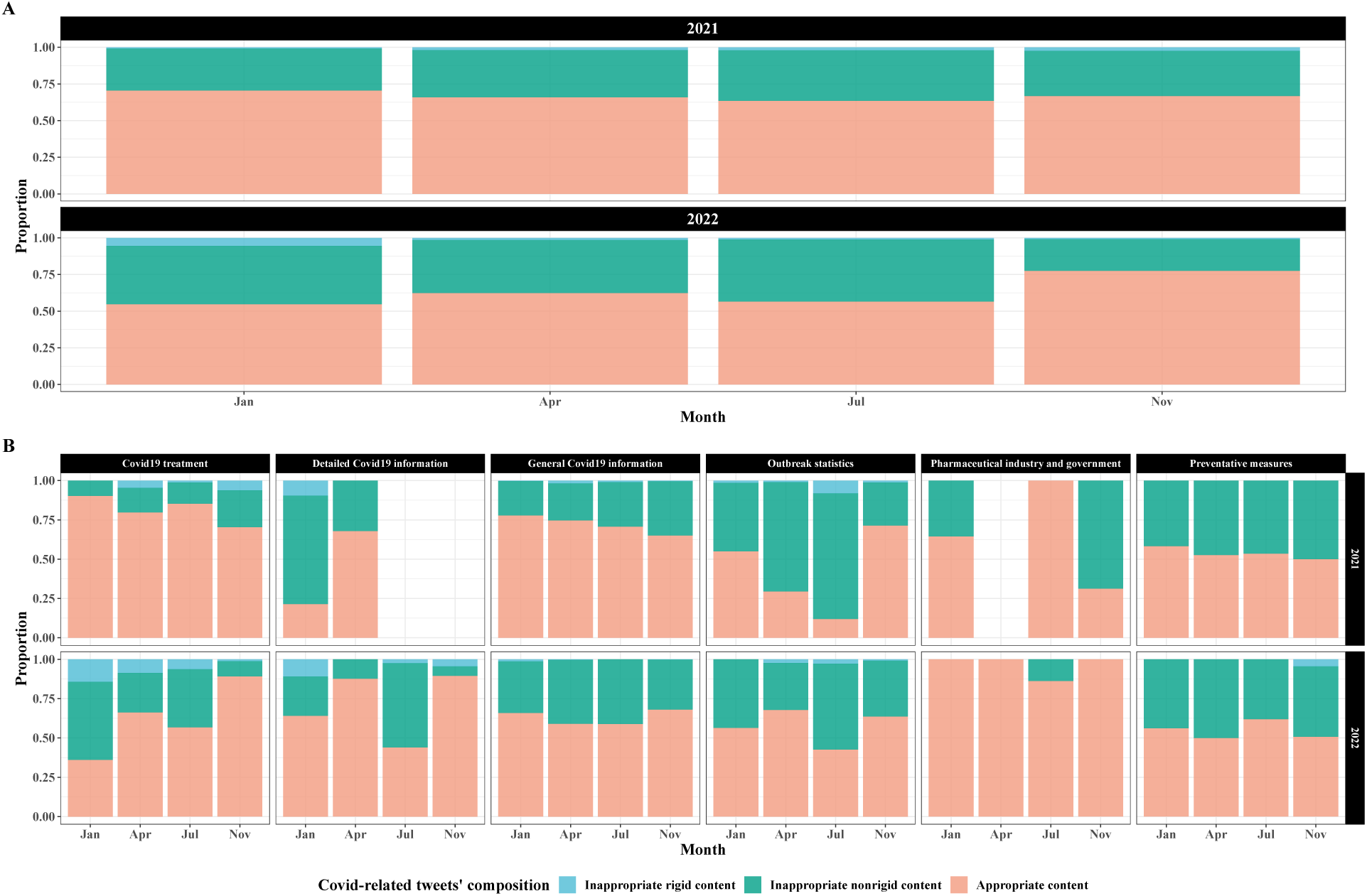
Tweet content composition based on the sentimental analysis. For each categorical level, we summarized the proportion of appropriate, inappropriate nonrigid, and inappropriate rigid content, and the detailed process can be found in Section 3.2.2. Appropriate content refers to content that expresses appropriate sentiments to promote hosts’ compliance, and inappropriate content is the opposite. Rigid refers to content with extremely inappropriate sentiments that may increase hosts’ rigidity or insensitivity to appropriate content. We collected Covid-related terms and their associated words with sentimental labels, and we summarized the proportion of appropriate, inappropriate nonrigid, and inappropriate rigid linkages for each Covid-related term and topic. Part A shows the content compositions for four months in the year 2021 and year 2022. Part B shows the content compositions for six Covid topics in four months of the year 2021 and year 2022.

## Appendix B: Feature and model selection

With the aim of building a robust, mechanistic, and relatively predictive framework that incorporates behavior and disease transmission, we chose social media as the primary driver of hosts’ behavioral change. Although media-driven behavioral change is a sound assumption to account for population-level behavioral change, what causes media change and how hosts respond to media change are two major problems that remain, and we want to address these two problems in this study.

First, we want to reduce the dimensionality of the problem and find out the most important predictors for Covid-related social media. We manually selected the most sensible predictors for the number of weekly new Covid-related tweets from the dataset (num): weekly new cases, weekly new deaths, the daily number of hospital patients averaged over each week, and weekly new vaccinations. As shown in Figure S3, based on the year 2021’s data, all four predictors are weekly correlated with the number of new weekly tweets, and there exists strong collinearity between new cases, new deaths, and hosp patients. In particular, new cases and hosp patients are most likely to contain repeated information. Indeed, VIF analysis also shows that hosp patients and new cases can be greatly explained by the remaining predictors, as shown in Table S2. Since new cases is somewhat more interpretable and easier to model, we dropped hosp patients, and VIF analysis after dropping hosp patients shows only a negligible amount of multi-collinearity.

**Figure S3:**
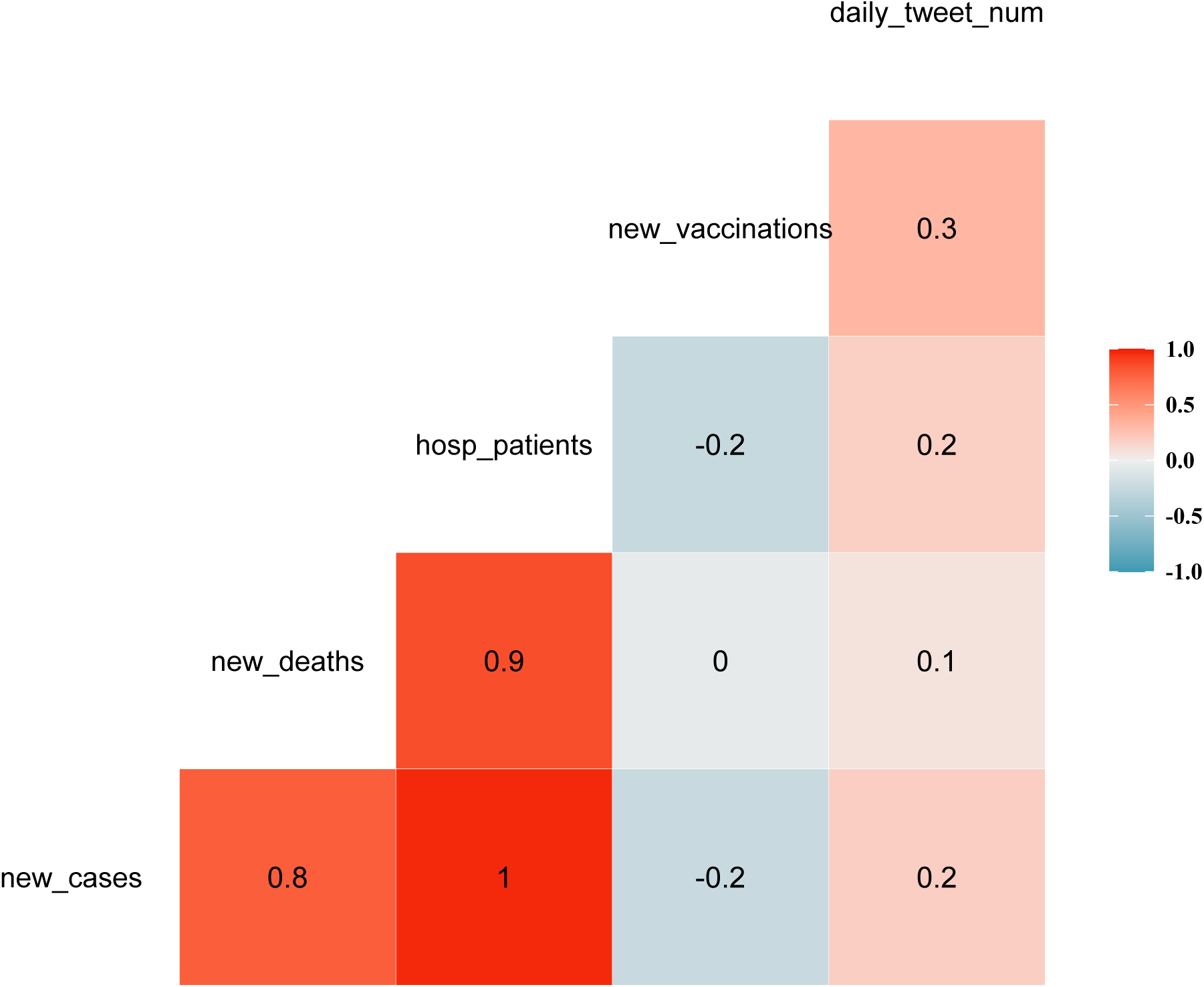
Correlations between different variables in the year 2021.

**Table S2:**
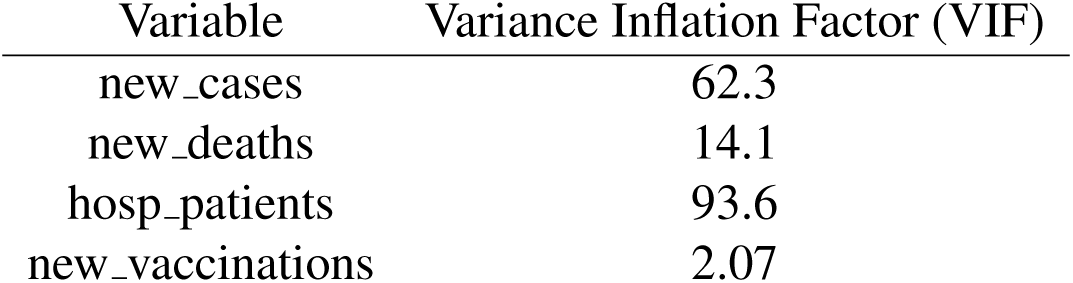
VIF table before eliminating variable hosp patients.

**Table S3:**
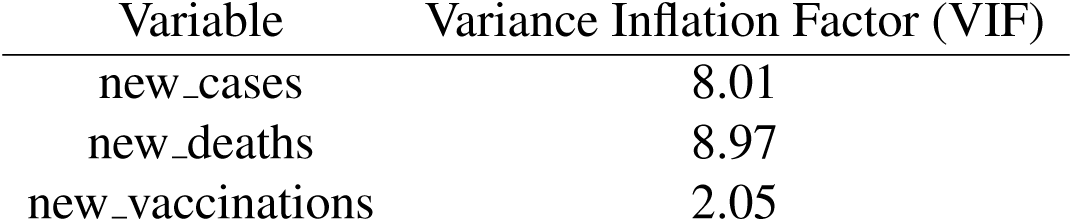
VIF table after eliminating variable hosp patients.

| Variable | Variance Inflation Factor (VIF) |
| --- | --- |
| new_cases | 8.01 |
| new_deaths | 8.97 |
| new_vaccinations | 2.05 |

For the rest of the predictors and all possible interactions between these predictors, we further narrowed them down with the best subset selection and cross-validation on the inner folds. As shown in S4, many models, those located on the flat curves, have similar test performances, suggesting that this framework is vulnerable to overfitting. In addition, due to the small test set size (8 observations), test error is highly variable, so if we simply choose the model with the minimum test error in each round, the selected models are likely going to be quite different from each other.

**Figure S4:**
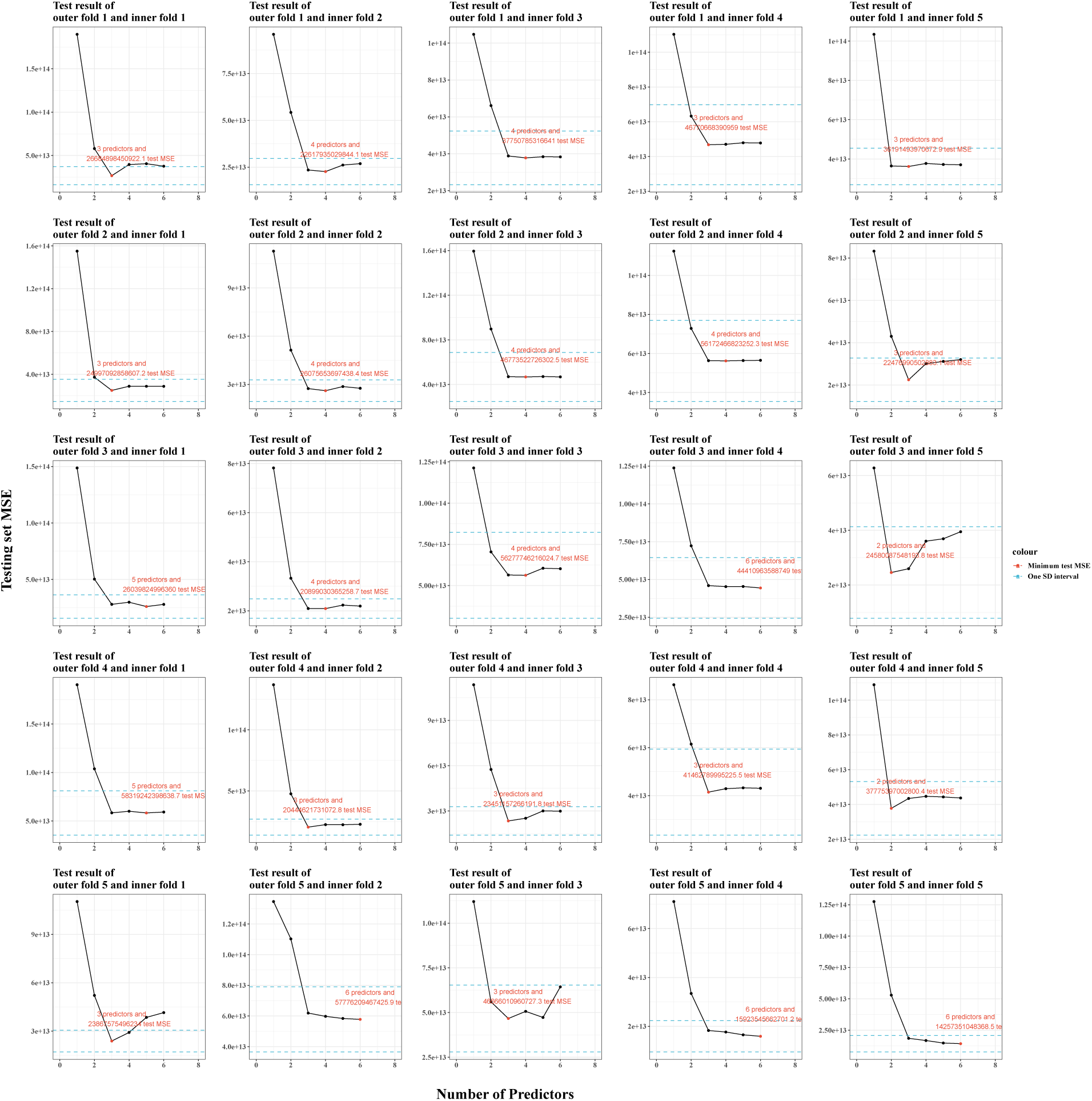
Best-subset feature selection on 25 inner folds of the cross-validation. We used the one-standard-error rule to select the optimal set of features in each round. The red dot in each subfigure shows the optimal feature set in that round.

Thus, to obtain a more robust model selection result, we choose the simplest model within the one-standard-error test-error interval of the best model (the one with the minimum test error) in each round. Since we have 5 outer folds and 5 inner folds, we got 25 rounds of model selection, and we summarized their results in Figure S5. In sum, new cases and new vaccinations are justified in all the selection trials, and their interaction is justified in most but not all the trials. However, for each outer fold (every five selection trials), there are at least one rounds in which the interaction is not justified. Again, we want the model that is the most parsimonious yet retains as much information as possible, so we finally pinned down new_cases and new_vaccinations as the predictors of new_media.

**Figure S5:**
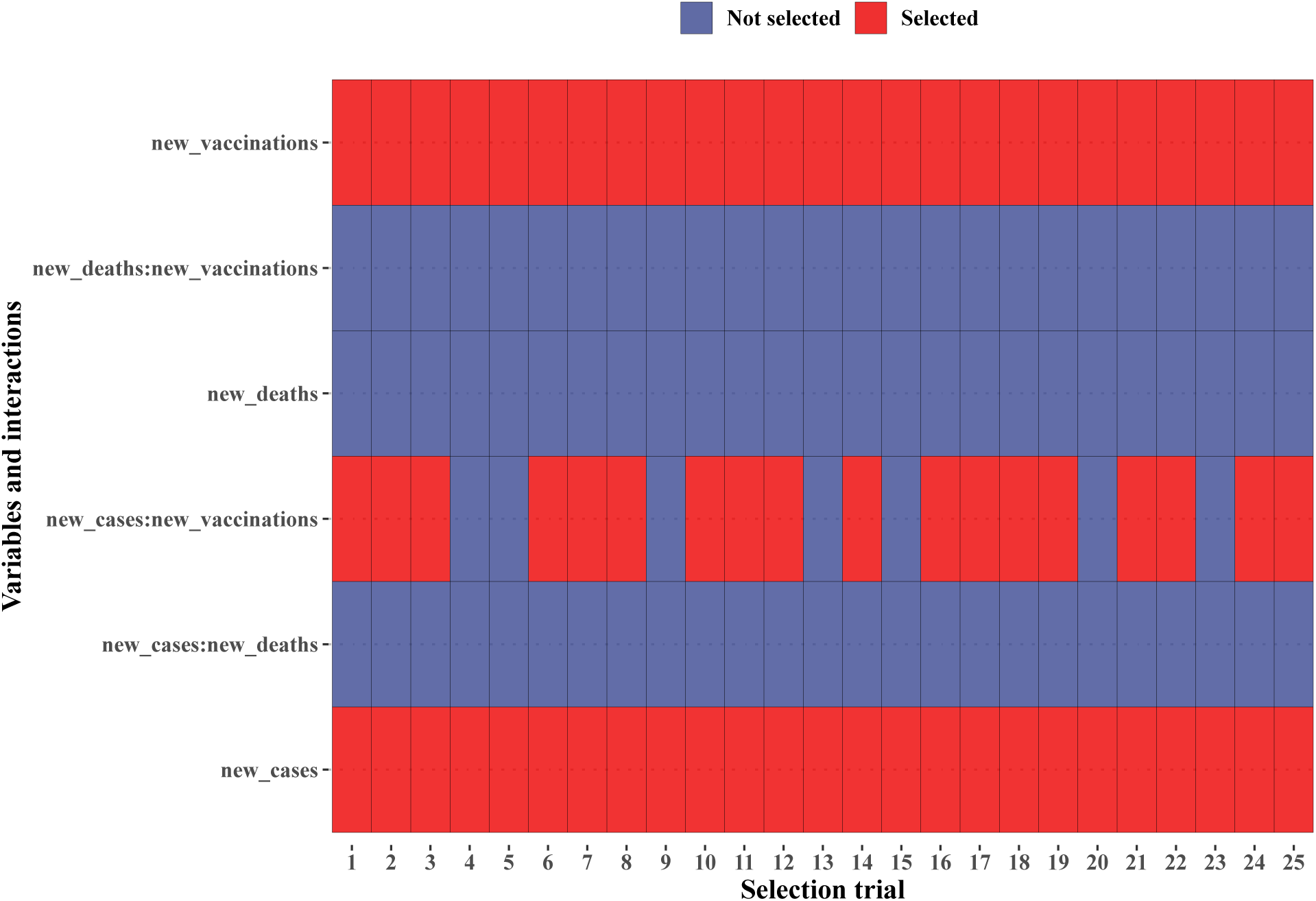
Feature selection result. One column represents one selection trial, with red and blue representing selected or unselected. Every five successive trials are operated on the same set of data, and for every five successive trials, we selected the parsimonious set of features.

While we can stop here and simply create a linear model to predict the weekly new media, we have not yet justified that each of these associations between the weekly new media and these two predictors is mostly linear. So, we square-transformed these two predictors and investigated their effect on the model performance. In addition, we considered “history” and “trend” as two other useful pieces of information to include as the production of media may have a time delay, and when the weekly new cases and/or weekly new vaccinations are increasing over weeks, media production will also accelerate. Thus, we incorporated two new encoded features, biweekly differences, into the model:

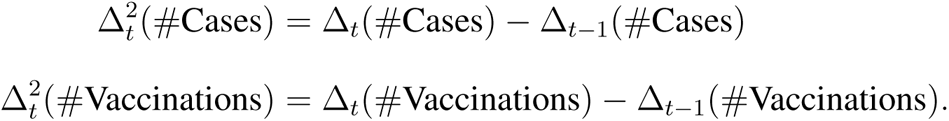

Now, the full model becomes

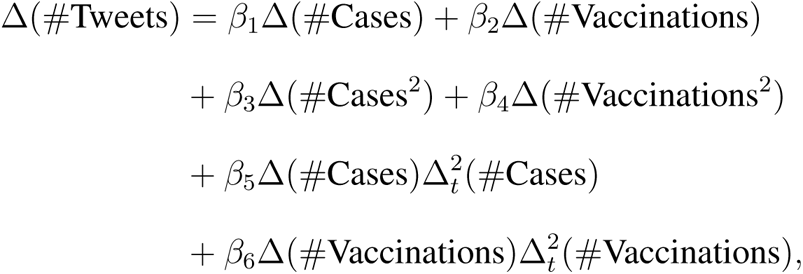

in which we modeled the curvatures of cases and vaccinations as effect modifiers of the weekly new cases and weekly new vaccinations. Then, we use the outer folds to select between several reduced models, and we still used the one-standard-error rule to select the final model. As shown in Table S4, the model with linear main effects and the interaction with the biweekly difference in the number of vaccinations has the lowest test CV. Within the one-standard-error interval of the best model, the simplest model is still the model with two linear main effects.

**Table S4:** Model selection results. One row represents one model and its 5-fold CV and the standard error of the CV. Models are ranked from the smallest CV to the largest CV, and we applied the one-standard-error rule to select the parsimonious model. + means the feature is in the model, and *−* means the feature is not in the model.

| Features | | | | | | CV, $10^{13}$ | se(CV), $10^{12}$ |
| --- | --- | --- | --- | --- | --- | --- | --- |
| $\Delta(\#Cases)$ | $\Delta(\#Vaccinations)$ | $\Delta(\#Cases^2)$ | $\Delta(\#Vaccinations^2)$ | $\Delta_t^2(\#Cases)$ | $\Delta_t^2(\#Vaccinations)$ | | |
| + | + | – | – | – | + | 5.49 | 5.93 |
| + | + | – | – | – | – | 5.71 | 8.79 |
| + | + | – | – | + | + | 5.72 | 7.21 |
| + | + | – | – | + | – | 5.92 | 9.20 |
| – | + | + | – | – | + | 7.01 | 5.78 |
| – | + | + | – | – | – | 7.07 | 6.23 |
| – | + | + | – | + | – | 7.97 | 11.5 |
| – | + | + | – | + | + | 8.16 | 15.1 |
| + | – | – | + | – | – | 8.64 | 21.4 |
| + | – | – | + | – | + | 8.75 | 16.3 |
| + | – | – | + | + | – | 9.29 | 23.5 |
| + | – | – | + | + | + | 9.45 | 20.0 |
| – | – | + | + | – | – | 14.0 | 22.0 |
| – | – | + | + | – | + | 14.9 | 18.9 |
| – | – | + | + | + | – | 16.5 | 36.0 |
| – | – | + | + | + | + | 17.8 | 40.1 |

In conclusion, we obtained the model from the above feature to predict the weekly new media.

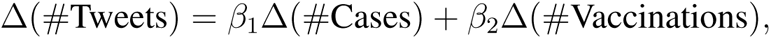

where Δ means the new observations in a week. Dividing Δ*t* to both sides and let Δ*t →* 0, we get

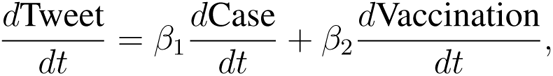

where *β*_1_ and *β*_2_ are positive. We can understand the model above easily: the rate of posting Covid-related media increases when the spread of the disease becomes faster and/or more and more hosts start to take vaccines.

## Appendix C: BMSIR parameter estimates and Bayesian inference with standard SIR model

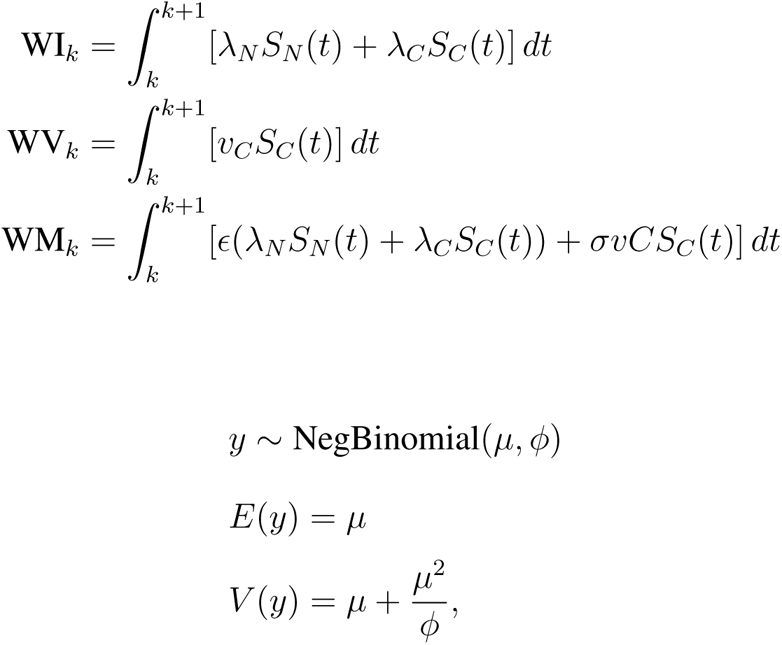

where 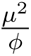 is the additional variance to the Poisson variance. For simplicity, we assumed that given the initial conditions and parameter values, weekly numbers are independent of each other, which is usually not the case for the stochastic process with significant process errors. Thus, the likelihood of the data is

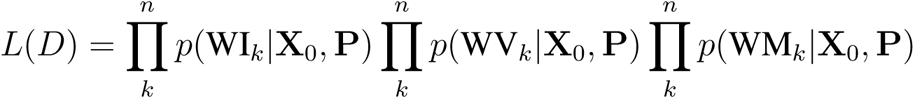

**Figure S6:**
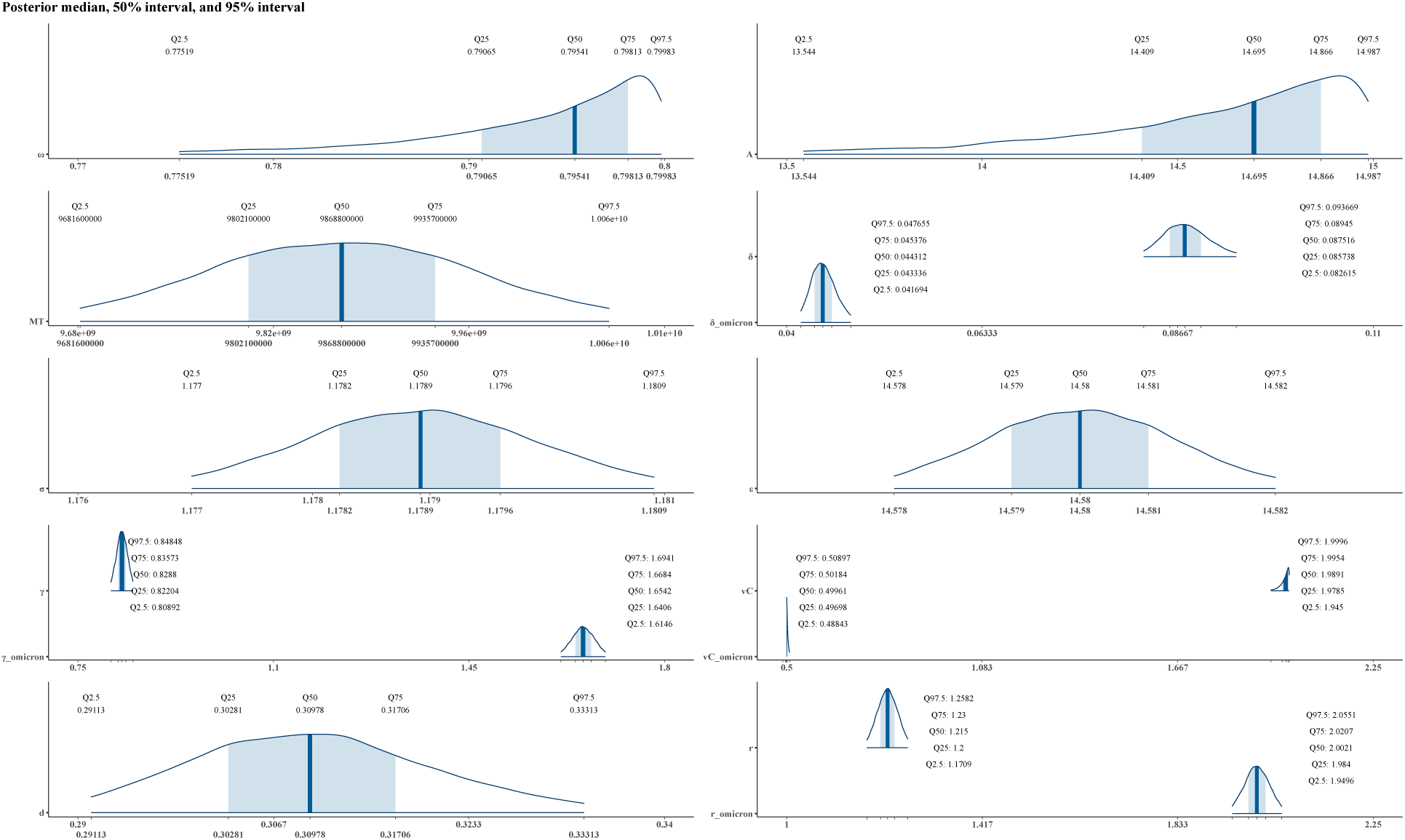
Posterior distributions of model parameters. Curves define the range of the 95% credible interval and the PDF of the distribution. Shaded areas are the 50% credible intervals. We also marked specific posterior percentiles in each subfigure.

**Figure S7:**
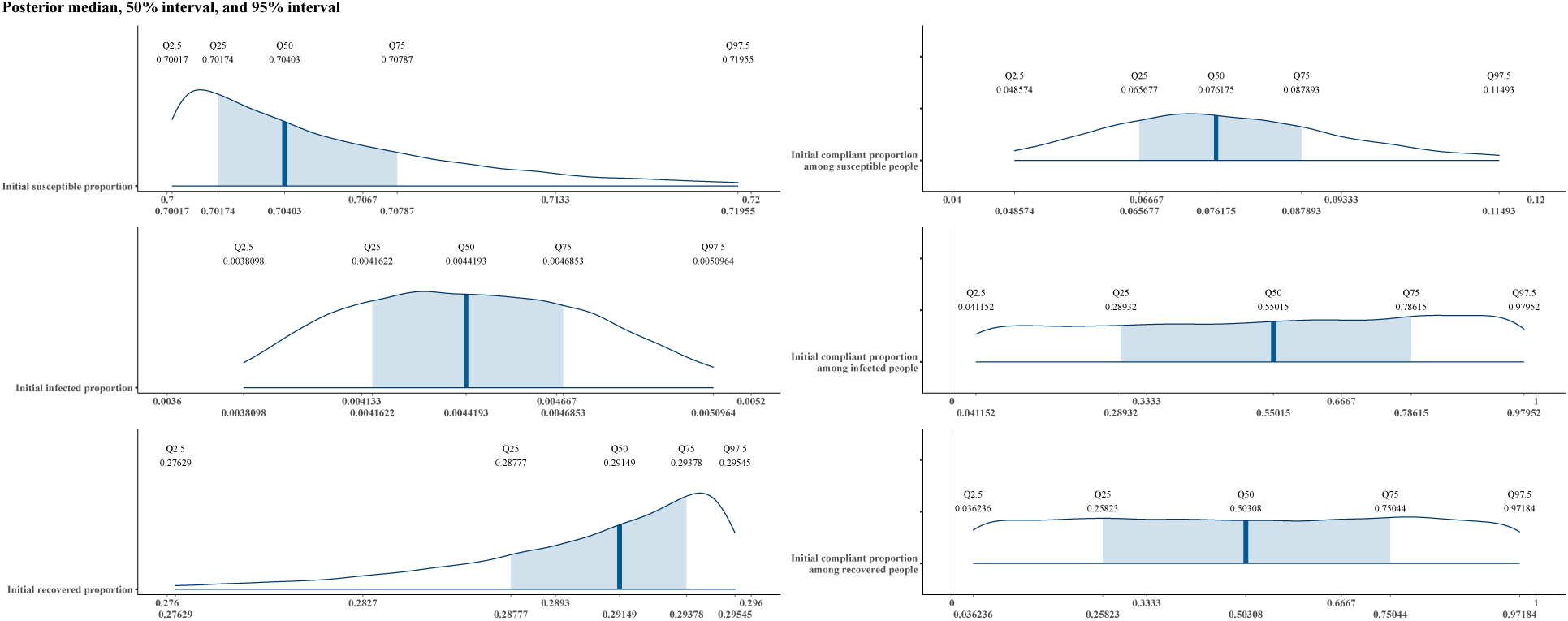
Posterior distributions of model initial conditions. Curves define the range of the 95% credible interval and the PDF of the distribution. Shaded areas are the 50% credible intervals. We also marked specific posterior percentiles in each subfigure.

**Figure S8:**
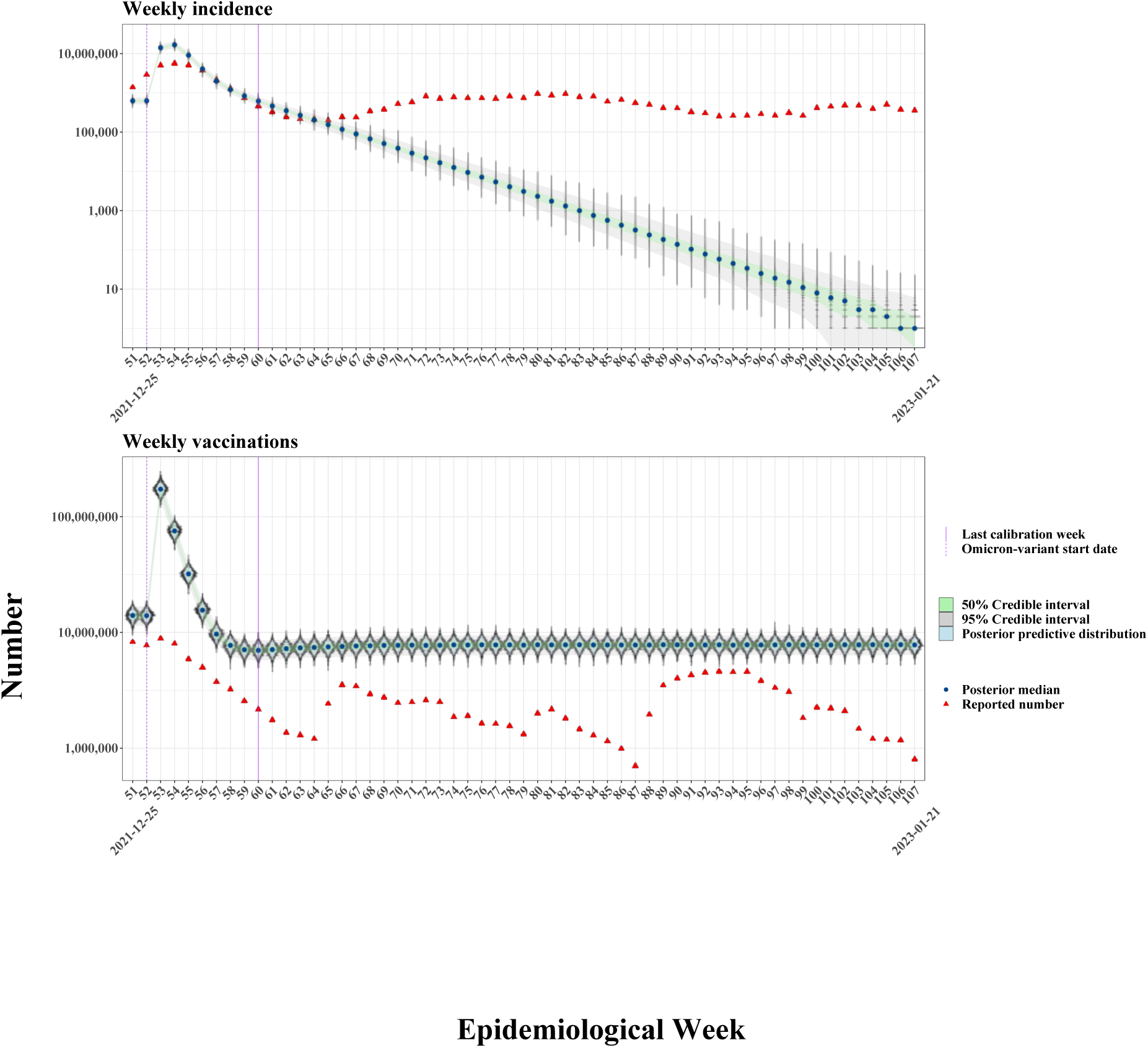
Bayesian posterior predictive distributions for the weekly data in the year 2021 using the standard SIR model. We used the same fitting procedures of the BMSIR model on the standard SIR model.

**Figure S9:**
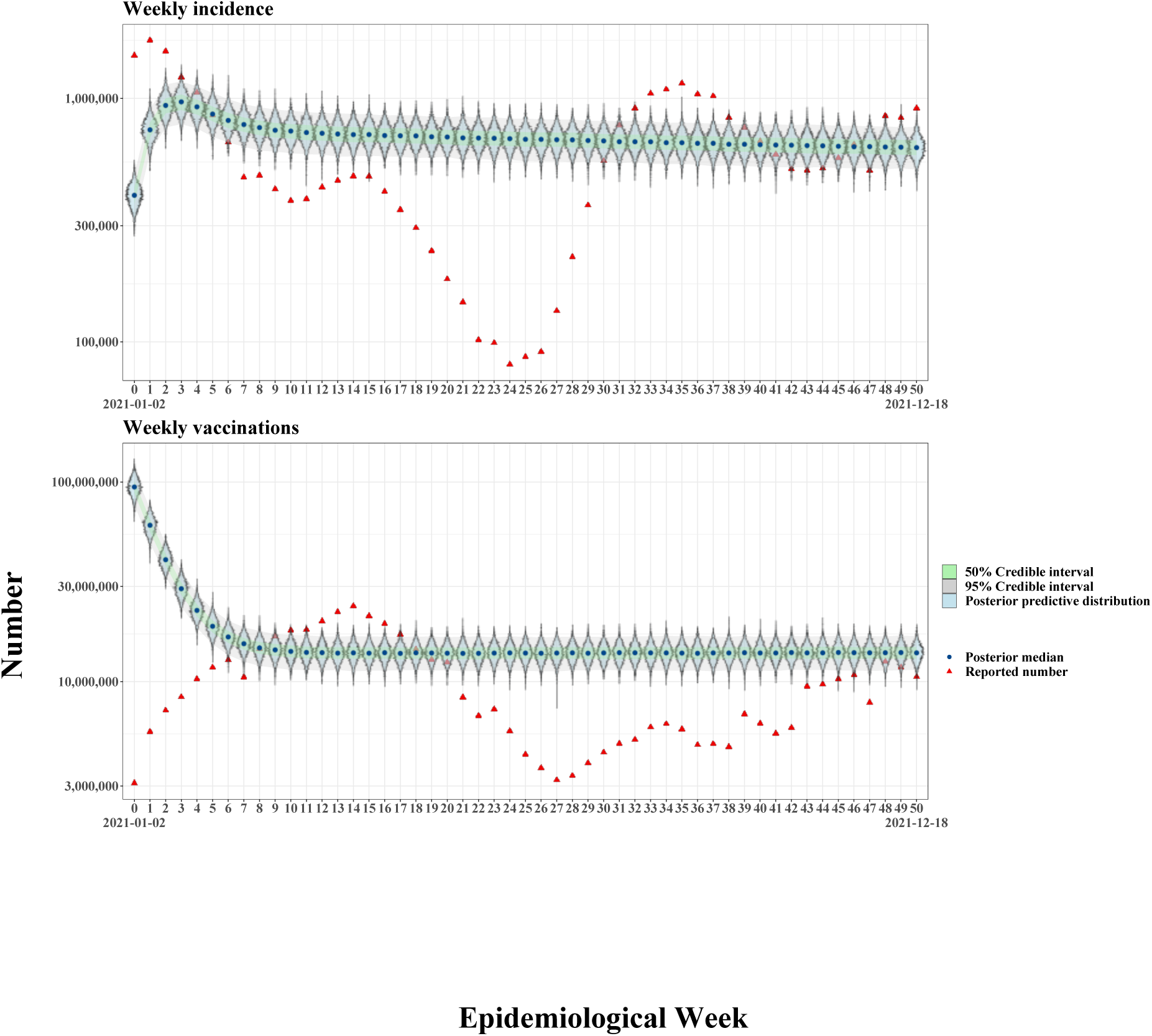
Bayesian posterior predictive distributions for the weekly data in the year 2021 and year 2022 using the standard SIR model. We used the same fitting procedures of the BMSIR model on the standard SIR model.

## References

[1] Grennan D. What is a Pandemic? Jama. 2019;321(9):910–0.

[2] Piret J, Boivin G. Pandemics throughout history. Frontiers in microbiology. 2021;11:631736.

[3] Schaller M. The behavioural immune system and the psychology of human sociality. Philosophical Transactions of the Royal Society B: Biological Sciences. 2011;366(1583):3418–26.

[4] Schaller M, Murray DR, Bangerter A. Implications of the behavioural immune system for social behaviour and human health in the modern world. Philosophical Transactions of the Royal Society B: Biological Sciences. 2015;370(1669):20140105.

[5] Güner HR, Hasanoğlu İ, Aktaş F. COVID-19: Prevention and control measures in community. Turkish Journal of medical sciences. 2020;50(9):571–7.

[6] Lau JT, Yang X, Tsui H, Pang E. SARS related preventive and risk behaviours practised by Hong Kong-mainland China cross border travellers during the outbreak of the SARS epidemic in Hong Kong. Journal of epidemiology & community health. 2004;58(12):988–96.

[7] Beutels P, Jia N, Zhou QY, Smith R, Cao WC, De Vlas SJ. The economic impact of SARS in Beijing, China. Tropical Medicine & International Health. 2009;14:85–91.

[8] Jones JH, Salathé M. Early assessment of anxiety and behavioral response to novel swine-origin influenza A (H1N1). PLoS one. 2009;4(12):e8032.

[9] Rubin GJ, Amlôt R, Page L, Wessely S. Public perceptions, anxiety, and behaviour change in relation to the swine flu outbreak: cross sectional telephone survey. Bmj. 2009;339.

[10] Al-Dmour H, Masa’deh R, Salman A, Abuhashesh M, Al-Dmour R. Influence of social media platforms on public health protection against the COVID-19 pandemic via the mediating effects of public health awareness and behavioral changes: integrated model. Journal of medical Internet research. 2020;22(8):e19996.

[11] Saud M, Mashud M, Ida R. Usage of social media during the pandemic: Seeking support and awareness about COVID-19 through social media platforms. Journal of Public Affairs. 2020;20(4):e2417.

[12] Tasnim S, Hossain MM, Mazumder H. Impact of rumors and misinformation on COVID-19 in social media. Journal of preventive medicine and public health. 2020;53(3):171–4.

[13] Verelst F, Willem L, Beutels P. Behavioural change models for infectious disease transmission: a systematic review (2010–2015). Journal of The Royal Society Interface. 2016;13(125):20160820.

[14] Fard LAN, Starnini M, Tizzoni M. Modeling adaptive forward-looking behavior in epidemics on networks. arXiv preprint arXiv:230104947. 2023.

[15] Berestycki H, Desjardins B, Weitz JS, Oury JM. Epidemic modeling with heterogeneity and social diffusion. Journal of Mathematical Biology. 2023;86(4):60.

[16] Clauß K, Kuehn C. Self-adapting infectious dynamics on random networks. arXiv preprint arXiv:220316949. 2022.

[17] Joseph JS, Zawawi JWM, Ahmad AH. The Effect of Gain Versus Loss Framing of Covid-19 Online News on Preventive Behavior. International Journal of Academic Research in Business and Social Sciences. 2021.

[18] Qian X, Xue J, Ukkusuri SV. Modeling disease spreading with adaptive behavior considering local and global information dissemination. arXiv preprint arXiv:200810853. 2020.

[19] Long Y. Spread and interaction of epidemics and information on adaptive social networks. The College of William and Mary; 2015.

[20] Abdulkareem SA, Augustijn EW, Mustafa YT, Filatova T. Intelligent judgements over health risks in a spatial agent-based model. International journal of health geographics. 2018;17(1):1–19.

[21] Zhang H, Small M, Fu X, Sun G, Wang B. Modeling the influence of information on the co-evolution of contact networks and the dynamics of infectious diseases. Physica D: Nonlinear Phenomena. 2012;241(18):1512–7.

[22] Yuan X, Xue Y, Liu M. Analysis of an epidemic model with awareness programs by media on complex networks. Chaos, Solitons & Fractals. 2013;48:1–11.

[23] XU Z, ZU Z, XU Q, Zhang W, Liu J, Zheng T. Effects of adaptive behavior on spreading dynamics of epidemics in structured populations. Military Medical Sciences. 2014:129–34.

[24] Funk S, Gilad E, Jansen VA. Endemic disease, awareness, and local behavioural response. Journal of theoretical biology. 2010;264(2):501–9.

[25] Misra A, Sharma A, Singh V. Effect of awareness programs in controlling the prevalence of an epidemic with time delay. Journal of Biological Systems. 2011;19(02):389–402.

[26] Misra A, Sharma A, Shukla J. Stability analysis and optimal control of an epidemic model with awareness programs by media. Biosystems. 2015;138:53–62.

[27] Samanta S, Rana S, Sharma A, Misra AK, Chattopadhyay J. Effect of awareness programs by media on the epidemic outbreaks: A mathematical model. Applied Mathematics and Computation. 2013;219(12):6965–77.

[28] Agaba G, Kyrychko Y, Blyuss K. Mathematical model for the impact of awareness on the dynamics of infectious diseases. Mathematical biosciences. 2017;286:22–30.

[29] Rai RK, Khajanchi S, Tiwari PK, Venturino E, Misra AK. Impact of social media advertisements on the transmission dynamics of COVID-19 pandemic in India. Journal of Applied Mathematics and Computing. 2022:1–26.

[30] Koutou O, Sangaré B, et al. Mathematical analysis of the impact of the media cover-age in mitigating the outbreak of COVID-19. Mathematics and Computers in Simulation. 2023;205:600–18.

[31] Guo J, Wang A, Zhou W, Gong Y, Smith SR. Discrete epidemic modelling of COVID-19 transmission in Shaanxi Province with media reporting and imported cases. Mathematical Biosciences and Engineering. 2022;19(2):1388–410.

[32] Tiwari PK, Rai RK, Khajanchi S, Gupta RK, Misra AK. Dynamics of coronavirus pandemic: effects of community awareness and global information campaigns. The European Physical Journal Plus. 2021;136(10):994.

[33] Chen E, Lerman K, Ferrara E. Tracking social media discourse about the covid-19 pandemic: Development of a public coronavirus twitter data set. JMIR Public Health and Surveillance. 2020;6(2):e19273.

[34] Banda JM, Tekumalla R, Wang G, Yu J, Liu T, Ding Y, et al. A large-scale COVID-19 Twitter chatter dataset for open scientific research—an international collaboration. Epidemiologia. 2021;2(3):315–24.

[35] Zhang L, Wang S, Liu B. Deep learning for sentiment analysis: A survey. Wiley Interdisciplinary Reviews: Data Mining and Knowledge Discovery. 2018;8(4):e1253.

[36] Mohammad S, Turney P. Emotions evoked by common words and phrases: Using mechanical turk to create an emotion lexicon. In: Proceedings of the NAACL HLT 2010 workshop on computational approaches to analysis and generation of emotion in text; 2010. p. 26–34.

[37] Mohammad SM, Turney PD. Crowdsourcing a word–emotion association lexicon. Computational intelligence. 2013;29(3):436–65.

[38] Lavine JS, Bjornstad ON, Antia R. Immunological characteristics govern the transition of COVID-19 to endemicity. Science. 2021;371(6530):741-5.

[39] Funk S, Gilad E, Watkins C, Jansen VA. The spread of awareness and its impact on epidemic outbreaks. Proceedings of the National Academy of Sciences. 2009;106(16):6872–7.

